# Plasma neurofilament light in behavioural variant frontotemporal dementia compared to mood and psychotic disorders

**DOI:** 10.1101/2023.02.19.23286151

**Authors:** Dhamidhu Eratne, Matthew Kang, Charles Malpas, Steve Simpson-Yap, Courtney Lewis, Christa Dang, Jasleen Grewal, Amy Coe, Hannah Dobson, Michael Keem, Wei-Hsuan Chiu, Tomas Kalincik, Suyi Ooi, David Darby, Amy Brodtmann, Oskar Hansson, Shorena Janelidze, Kaj Blennow, Henrik Zetterberg, Adam Walker, Olivia Dean, Michael Berk, Cassandra Wannan, Christos Pantelis, Samantha Loi, Mark Walterfang, Samuel Berkovic, Alexander Santillo, Dennis Velakoulis, The MiND Study Group

## Abstract

**Objective:** Blood biomarkers of neuronal injury such as neurofilament light (NfL) are being intensively studied to improve diagnosis and treatment of neurodegenerative disorders, but gaps remain in its ability to assist in distinguishing neurodegenerative from primary psychiatric disorders (PPD) with overlapping clinical presentations that commonly cause diagnostic dilemmas. This study aimed to investigate plasma NfL in a range of PPDs, and the diagnostic utility of plasma NfL in differentiating PPD from behavioural variant frontotemporal dementia (bvFTD), a neurodegenerative disorder commonly misdiagnosed initially as PPD. Furthermore, improved understanding of NfL in a diverse range of PPDs, the role and performance of a large normative/reference data sets and models to facilitate precision interpretation of an individual’s levels, and the influence of covariates, will be critical for future research and clinical translation.

**Methods:** Plasma NfL was analysed using Single molecule array (Simoa) technology in major depressive disorder (MDD, n=42), bipolar affective disorder (BPAD, n=121), treatment-resistant schizophrenia (TRS, n=82), and bvFTD (n=22). Comparisons were made between the four clinical cohort groups, and the reference cohort (Control Group 2, n=1926, using generalised additive models for location, scale, and shape (GAMLSS), and age-matched controls (Control Group 1, n=96, using general linear models),

**Results:** Large differences were seen between bvFTD (mean NfL 34.9pg/mL) and all PPDs and controls (all <11pg/mL). Plasma NfL distinguished bvFTD from PPD with high accuracy; a 13.3pg/mLcut-off resulted in 86% sensitivity, 88% specificity. GAMLSS models using the large Control Group 2 performed equally to or outperformed models using local controls. An internet-based application was developed to provide individualised z-scores and percentiles based on this reference cohort, which can facilitate precision interpretation of an individual level. Slightly higher plasma NfL levels were found in BPAD, compared to both control groups, and compared to TRS.

**Conclusions:** This study adds further evident on the strong diagnostic utility of NfL to distinguish bvFTD from clinically relevant PPDs, and includes the largest cohort of BPAD to date. The finding of higher plasma NfL levels in the largest cohort of BPAD to date should prompt further investigation. Use of large reference cohorts and GAMLSS modelling may have important implications for future research and clinical translation. Studies are underway investigating clinical and diagnostic utility of plasma NfL and the serviceability of the internet-based application for diverse neurodegenerative and primary psychiatric conditions in real-world primary care and specialist clinical settings.

Table 1.

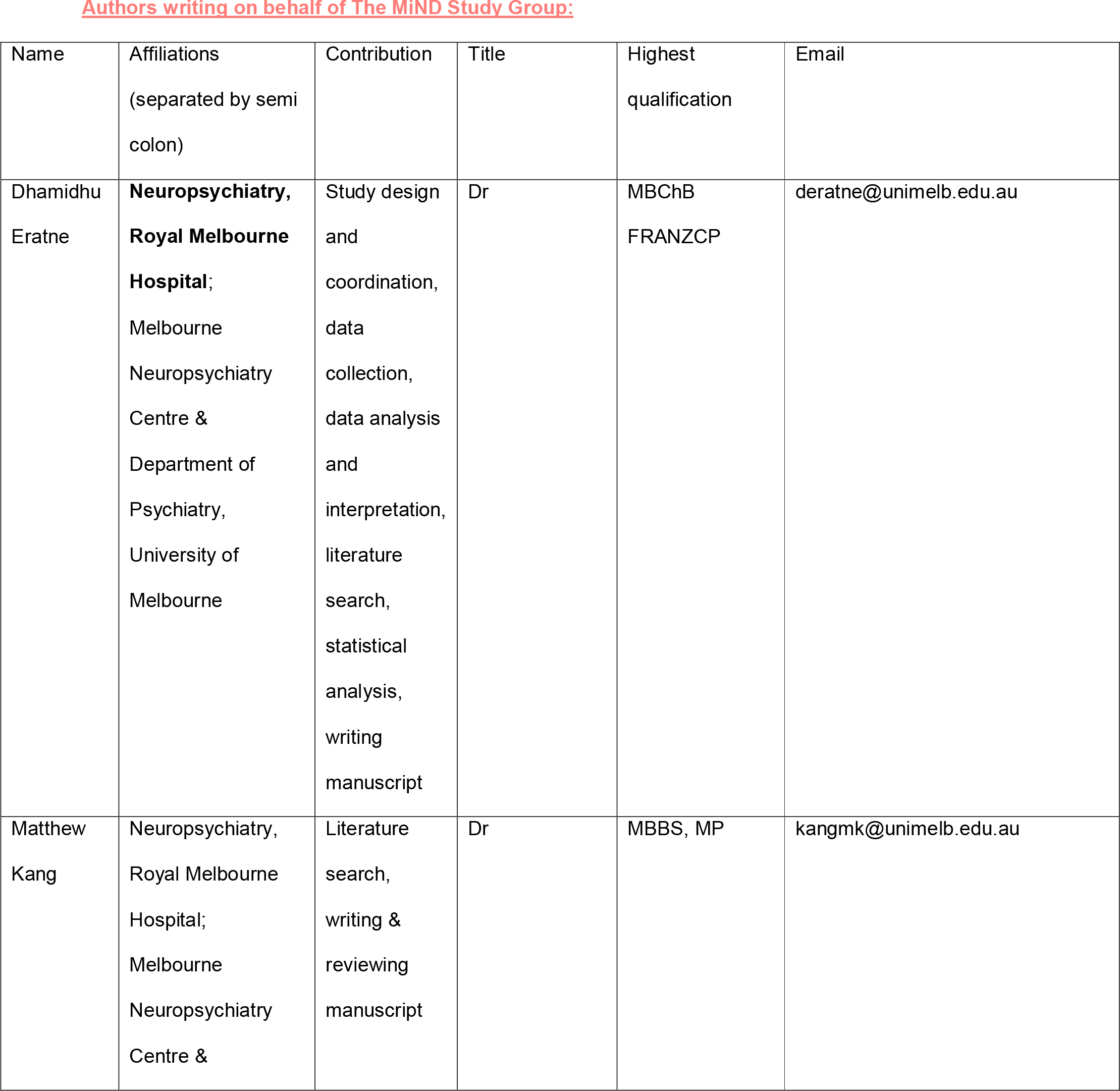

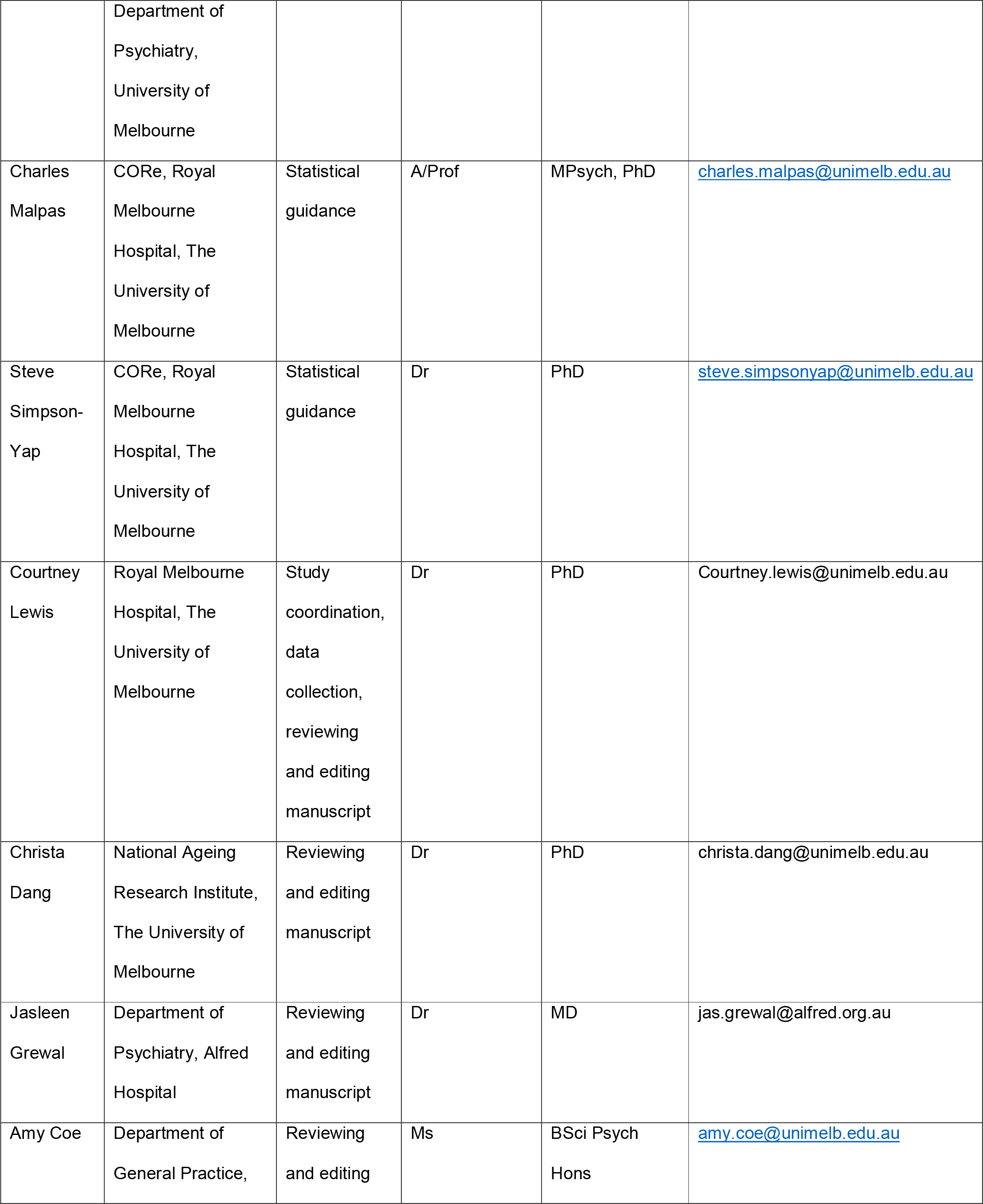

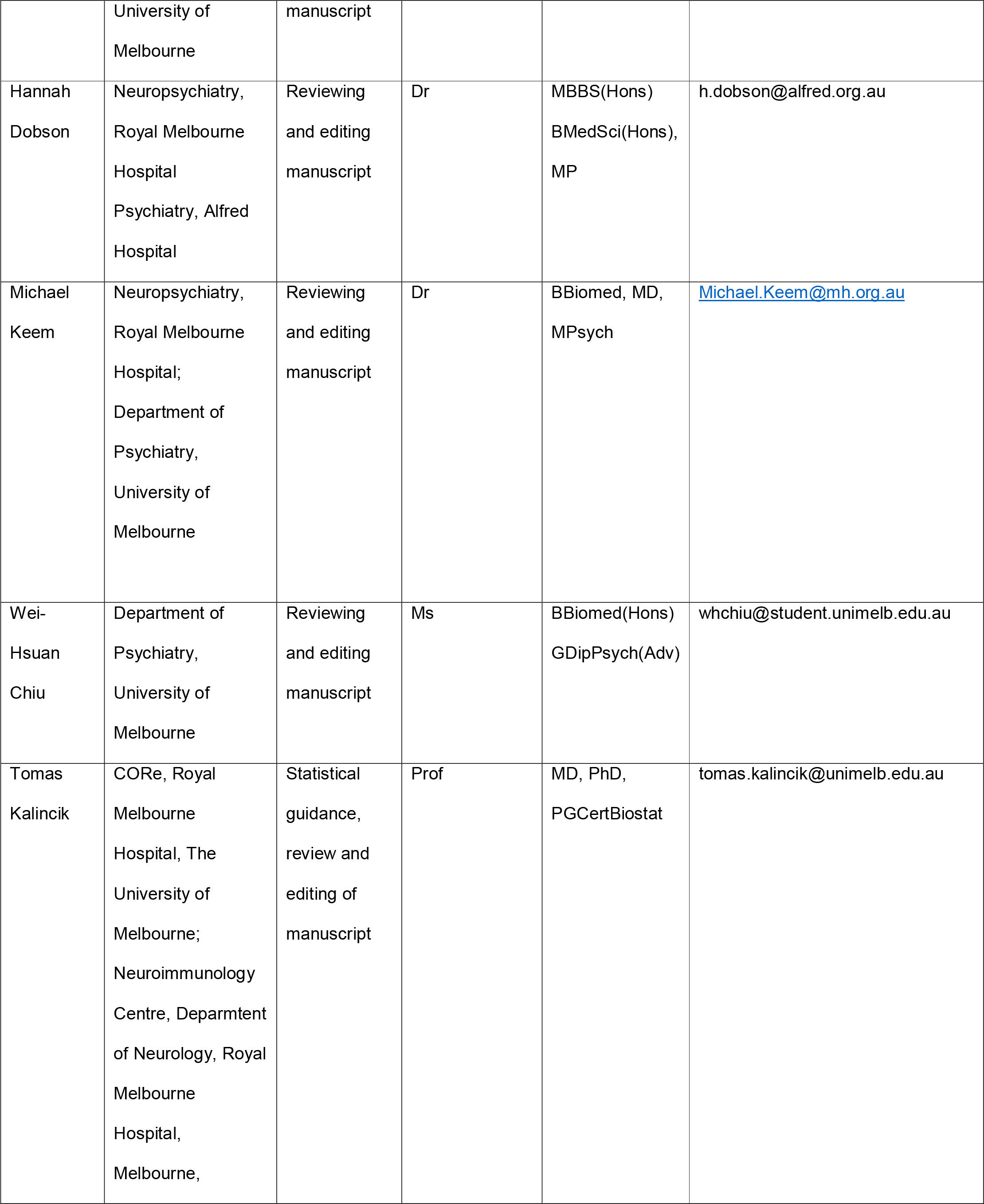

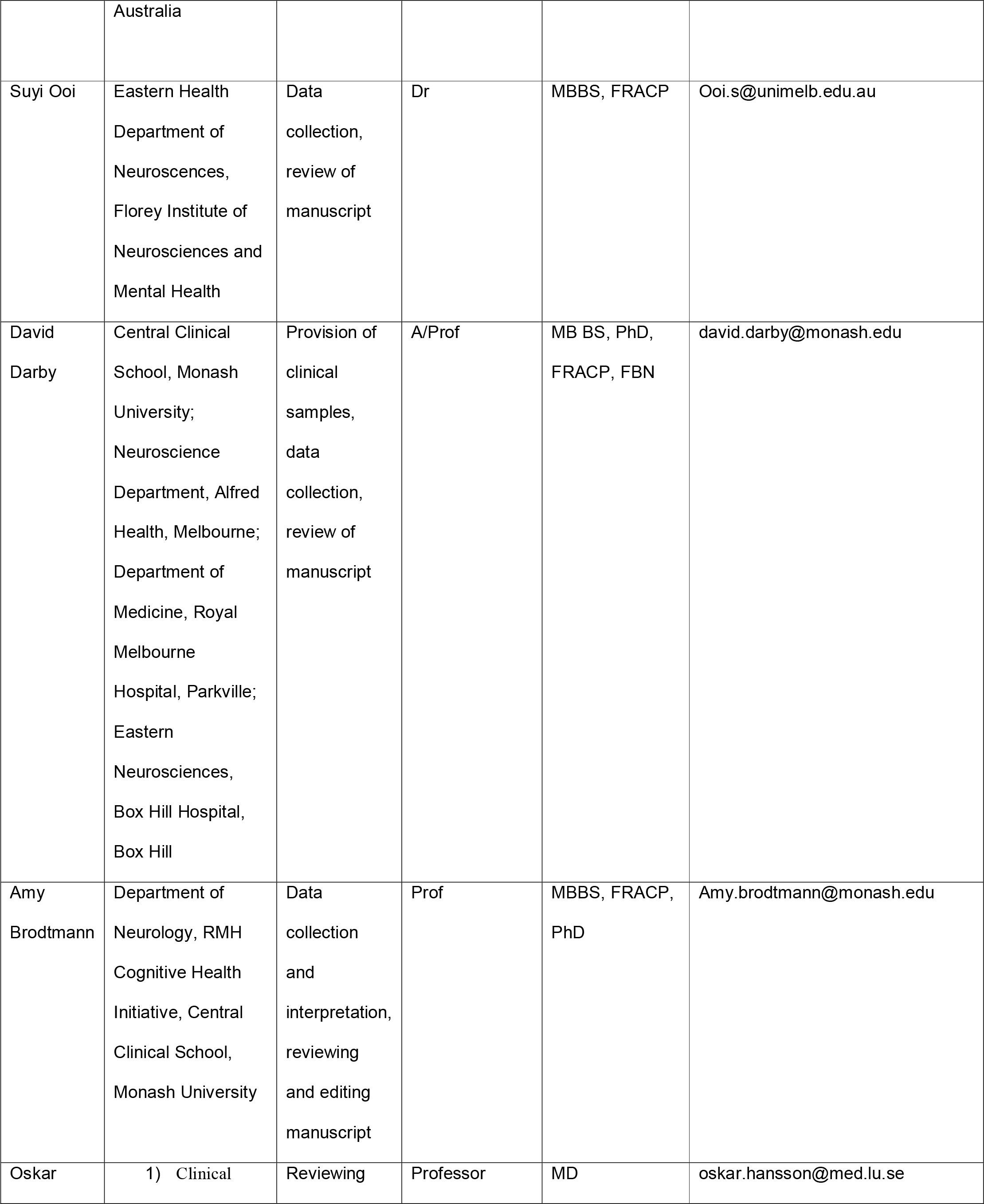

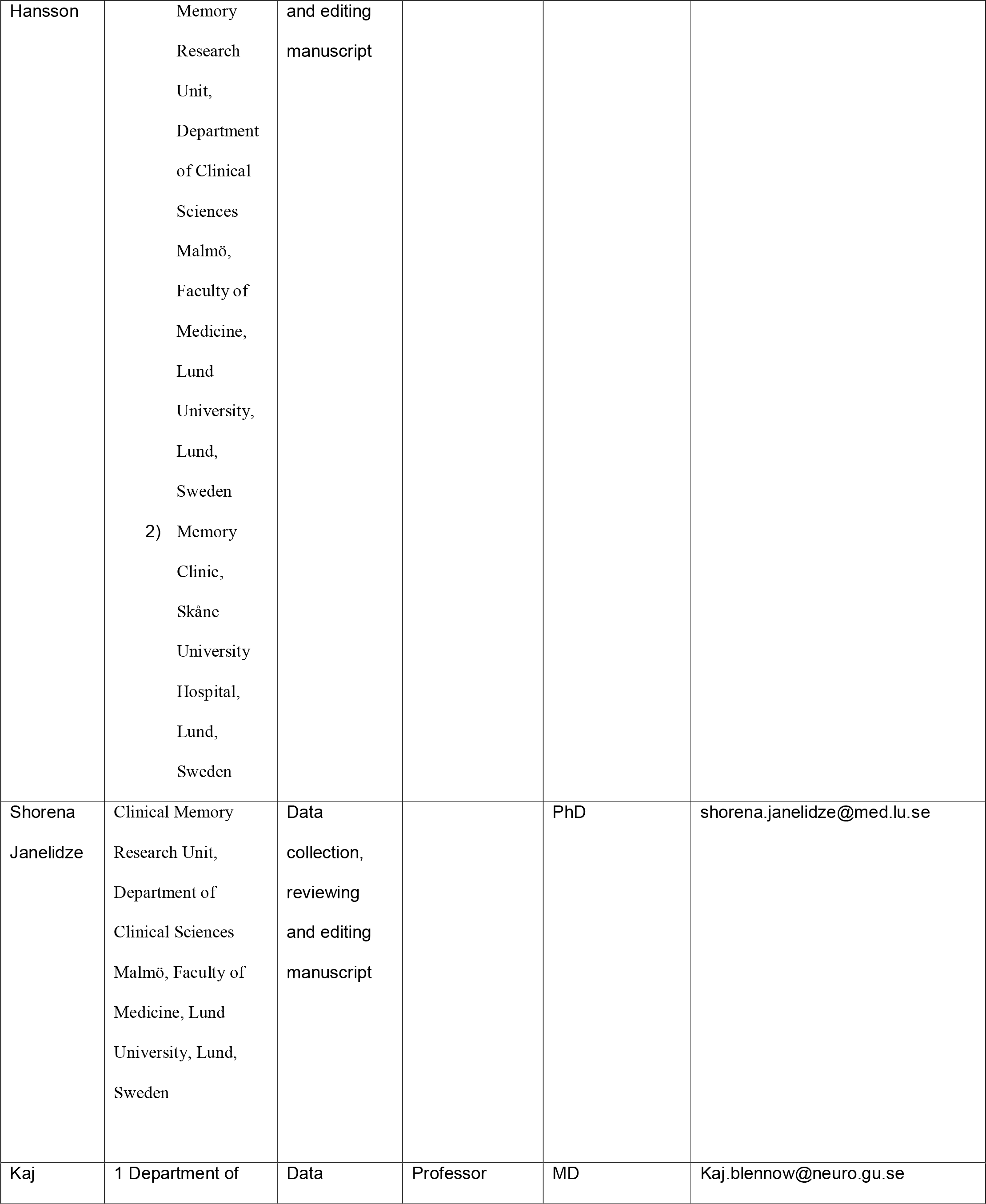

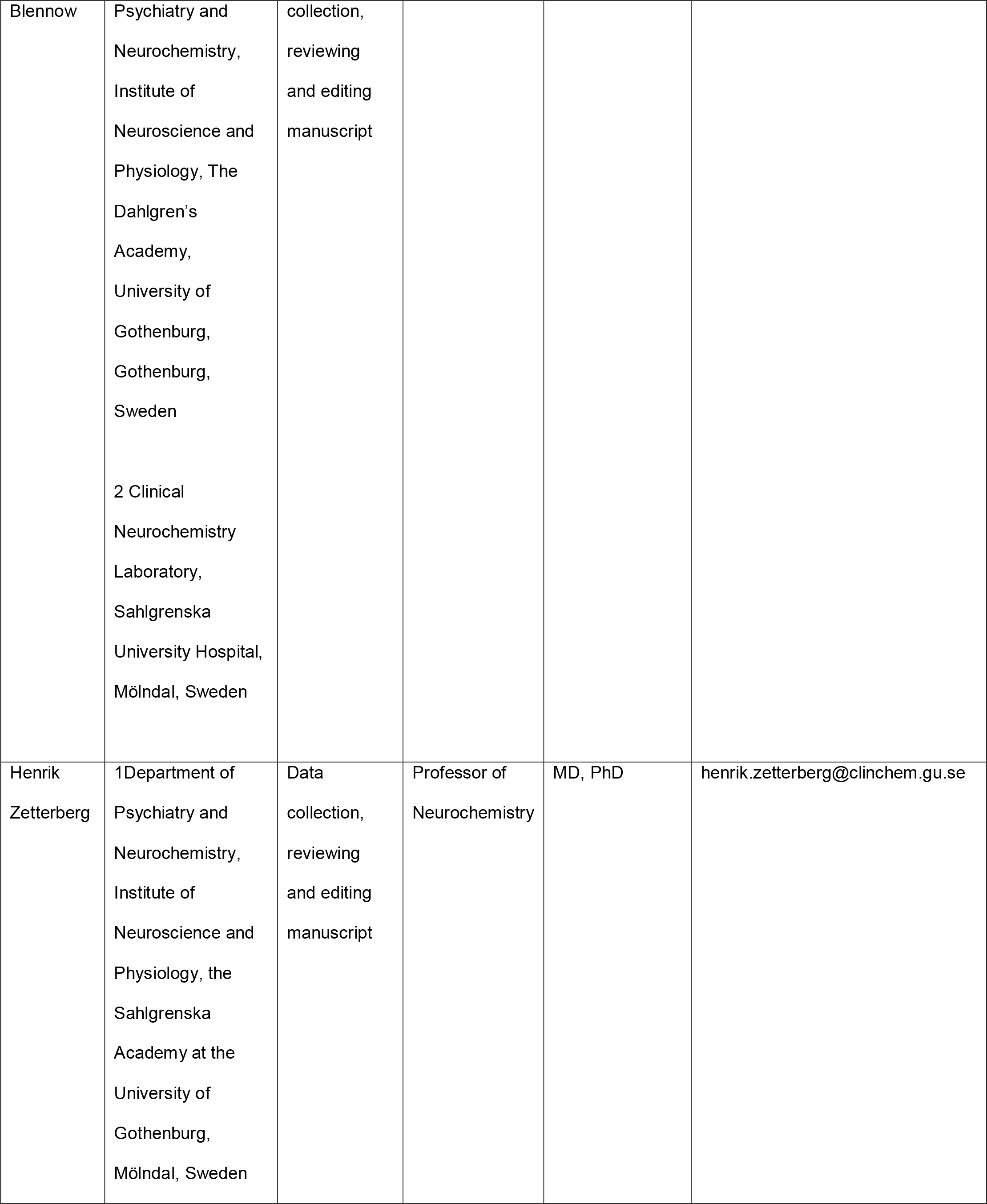

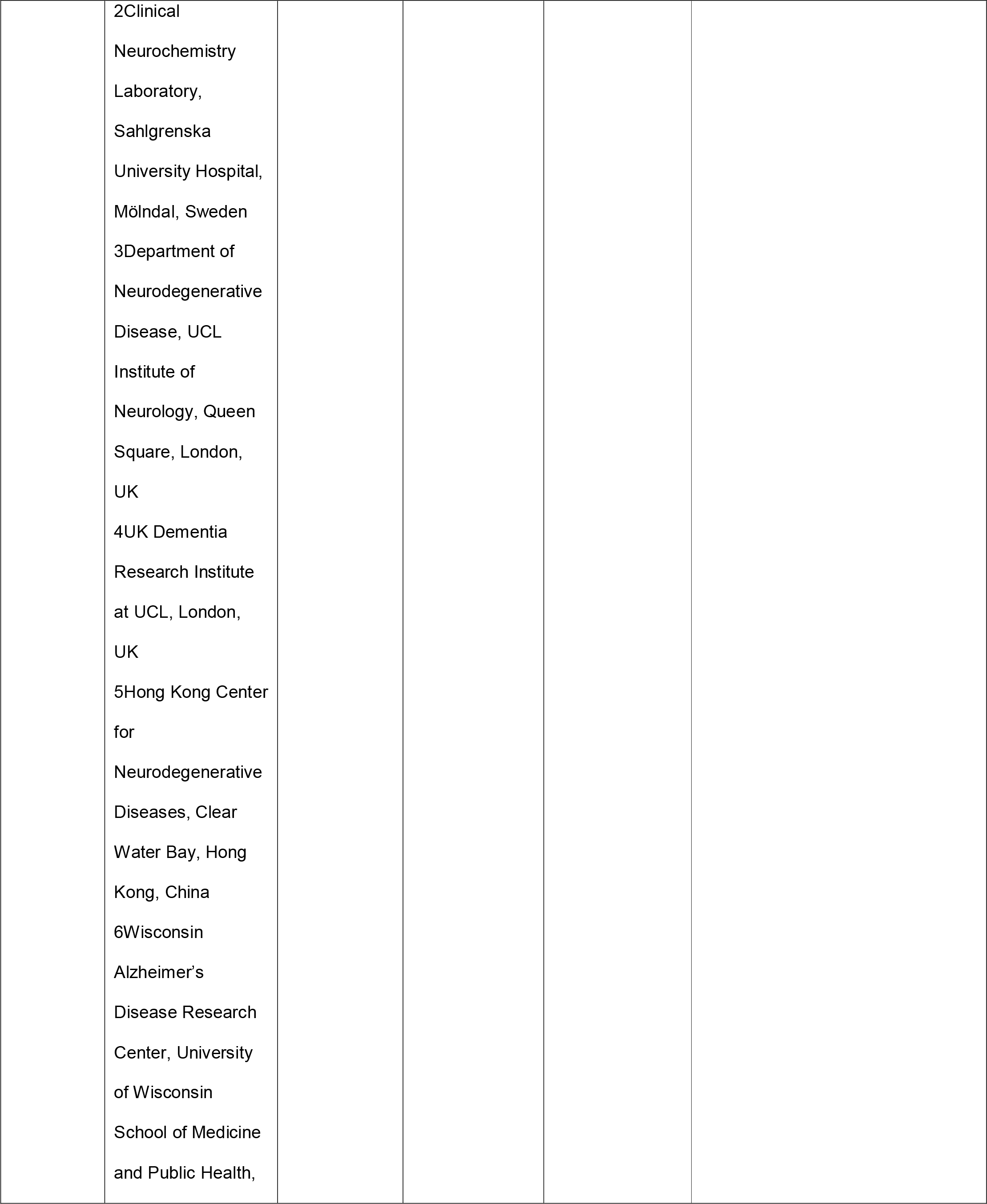

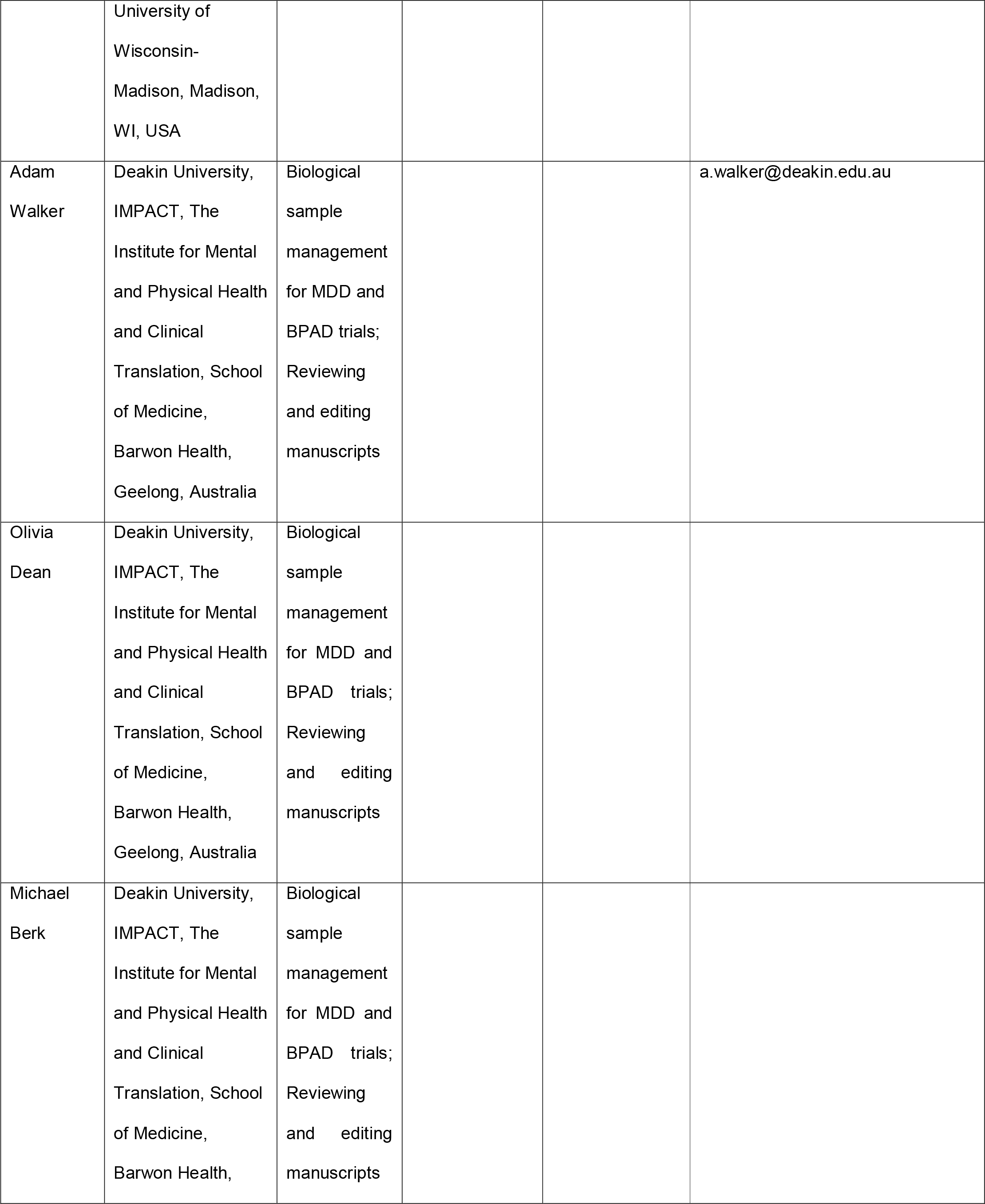

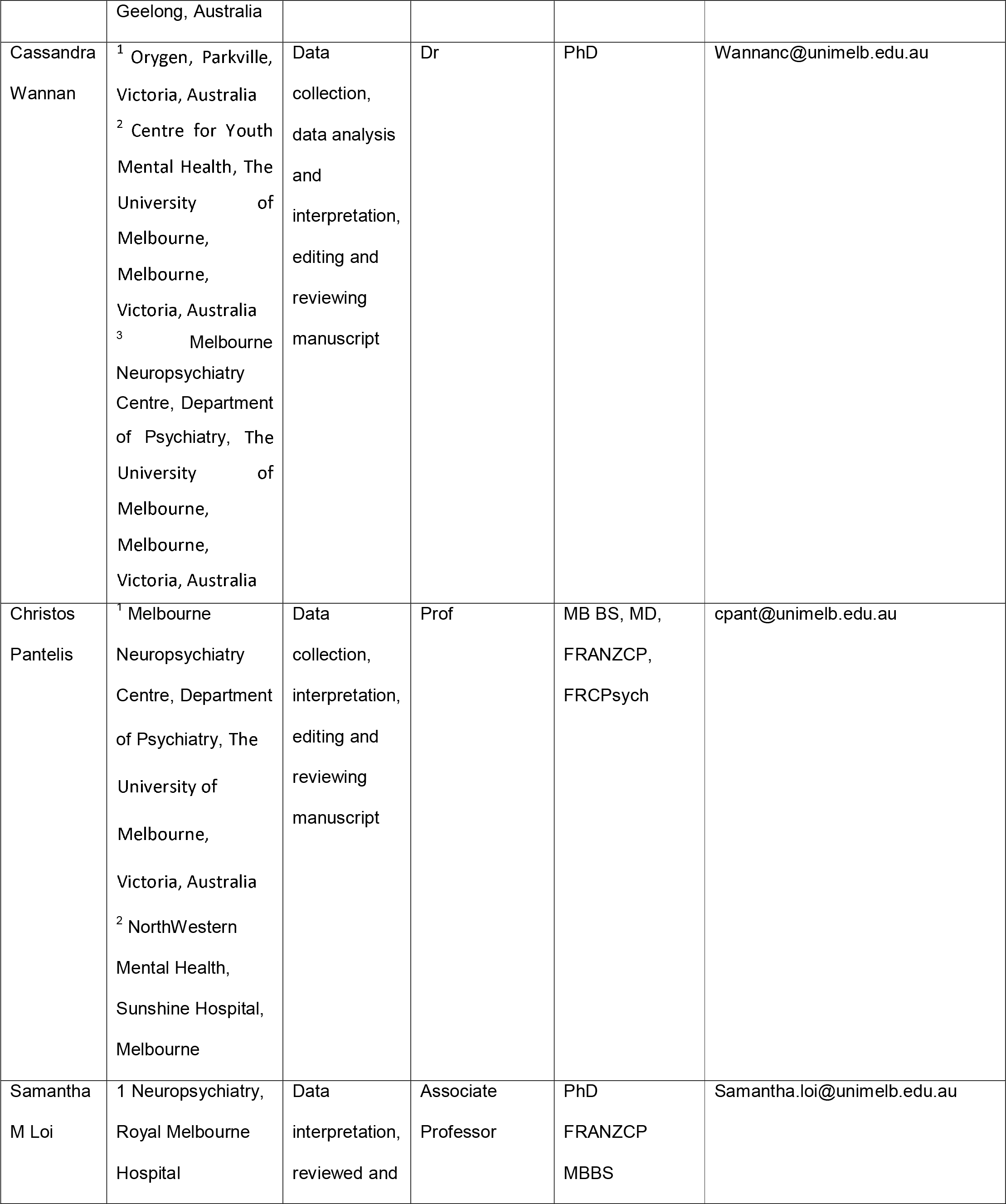

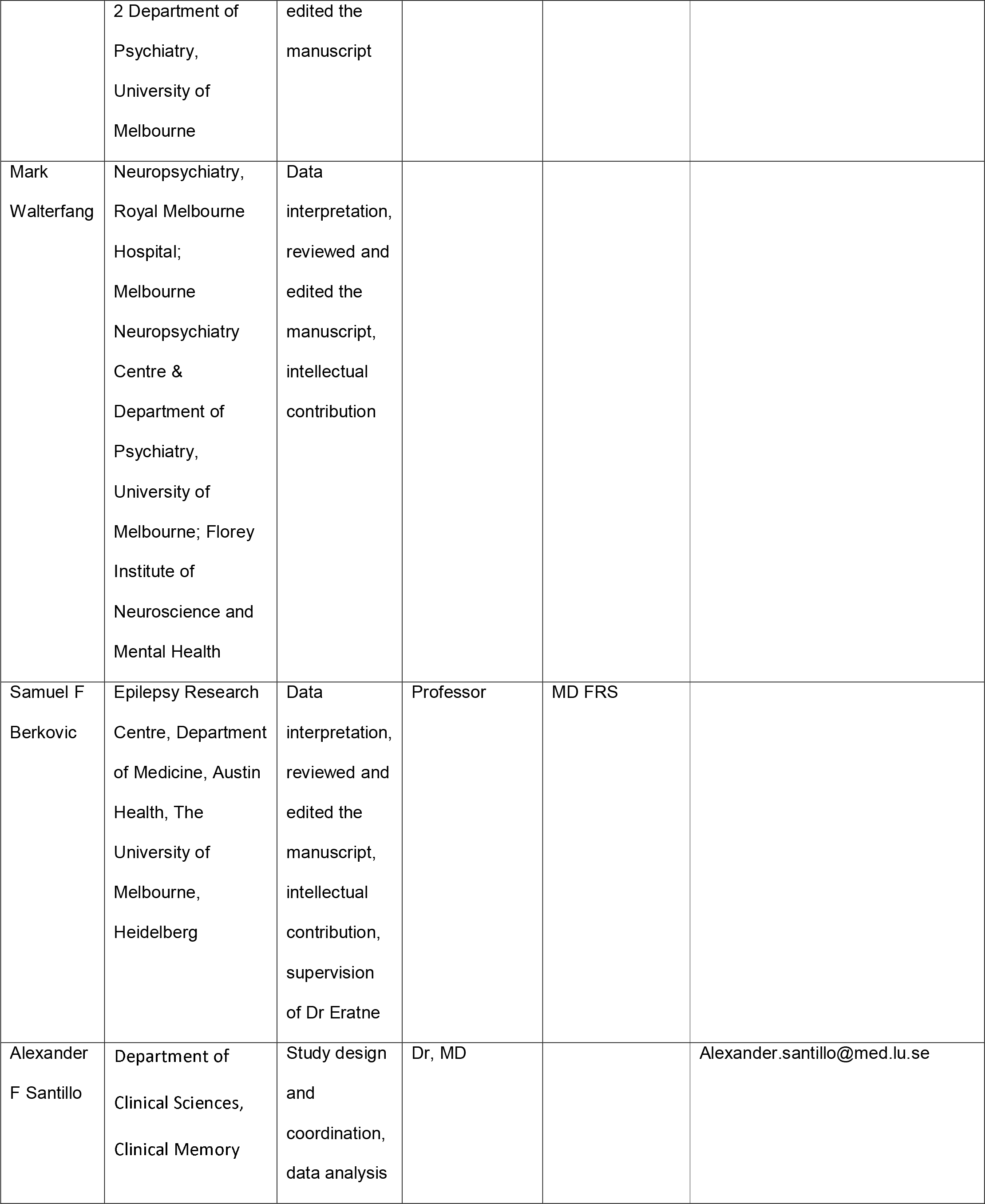

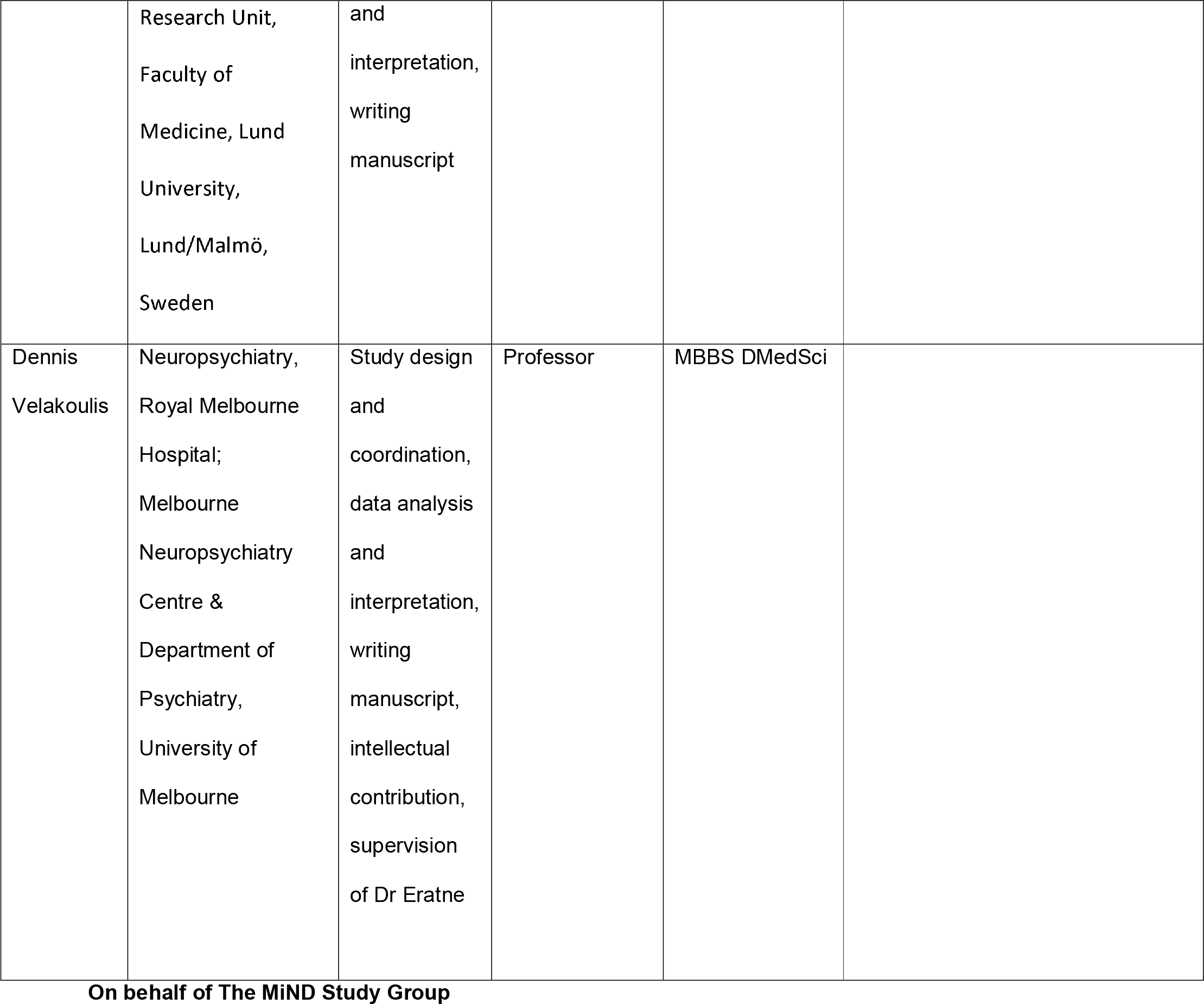

## INTRODUCTION

There has been a great deal of research on blood-based biomarkers for neurological and neurodegenerative conditions, with implications for clinical trials and clinical translation to improve early diagnosis (even pre-clinical diagnosis), care and treatment. In particular, neurofilament light chain (NfL) has been demonstrated to be a highly sensitive biomarker of neuronal injury in a diverse range of conditions (Bridel et al., 2019; Eratne et al., 2020; Eratne, Keem, et al., 2022; Eratne, Loi, et al., 2022; Gaetani et al., 2019, 2021; Khalil et al., 2018). NfL is of particular interest as a potential diagnostic biomarker, as it may help distinguish disorders with significant neuronal degeneration, from those without. Distinguishing neurodegenerative dementias from primary psychiatric disorders (PPD) is a frequent clinical diagnostic dilemma and one associated with uncertainty, misdiagnosis, and negative impacts for patients and healthcare systems (Kang et al., 2022; Loi et al., 2020; Tsoukra et al., 2021; Woolley et al., 2011).

One of the most challenging clinical distinctions, associated with some of the highest diagnostic uncertainty/instability and misdiagnoses, is distinguishing primary psychiatric disorders (PPD) from behavioural variant frontotemporal dementia (bvFTD), a neurodegenerative condition associated with variable personality, behavioural and psychiatric changes (Ducharme et al., 2020; Eratne, Keem, et al., 2022; Kang et al., 2022; Tsoukra et al., 2021). Previous studies have investigated blood and cerebrospinal fluid (CSF) NfL in bvFTD compared to PPD (Al Shweiki et al., 2019; Ashton et al., 2021; Katisko et al., 2020; Vijverberg et al., 2017), and in bvFTD and range of other neurodegenerative conditions compared to PPD and non-progressive/phenocopy syndromes (Eratne et al., 2020; Eratne, Keem, et al., 2022; Eratne, Loi, et al., 2022; Fourier et al., 2020), finding significantly elevated levels in neurodegenerative disorders including bvFTD compared to PPDs. There remain significant gaps in our understanding of NfL levels in large, well characterised cohorts of severe PPDs that can often present or ‘mimic’ conditions like bvFTD, and/or are associated themselves with cognitive and neuroimaging abnormalities.

None of the previously mentioned studies have used large normative samples, which would allow inferences at the individual patient level. There is increasing interest in the use of models derived from and comparisons to very large reference cohorts, to facilitate more precise interpretation of individual levels, improving upon and moving beyond coarse age-binned binary cut-offs (Benkert et al., 2022).

Improved understanding of models based on large reference cohorts, with a specific focus on utility in distinguishing bvFTD from PPD, will have additional important implications for future research and clinical translation.

Few studies to date have investigated blood NfL with a specific focus on PPD, and the findings have been mixed. Higher blood NfL levels were seen in MDD compared to controls in one study (Bavato et al., 2021), but not in another study (Ashton et al., 2021). Higher serum NfL levels were seen in bipolar depression (Aggio et al., 2022). Mixed findings have also been seen in psychotic disorders: some studies reported no differences between controls and schizophrenia (Al Shweiki et al., 2019; Bavato et al., 2021) and between controls and treatment-resistant clozapine-treated schizophrenia (Eratne et al., 2021) while one study reported slightly higher NfL levels in both schizophrenia and clozapine-treated patients compared to controls (Rodrigues-Amorim et al., 2020). A recent study found higher serum NfL levels in psychosis associated with anti-NMDA receptor encephalitis, compared to psychosis due to primary psychiatric illness, of particular relevance to patients in psychiatric settings (Guasp et al., 2022).

Improved understanding of the ranges and performance of NfL in a diverse range of PPDs, particularly those of clinical relevance given overlapping symptoms with neurodegenerative differential diagnoses, such as mood and psychotic disorders, will be critical for broad clinical translation in to diverse clinical settings.

**<u>Aim 1</u>** of this study was to compare plasma NfL levels in bvFTD to primary psychiatric conditions that can often appear like or ‘mimic’ bvFTD, such as bipolar affective disorder (BPAD), major depressive disorder (MDD), and schizophrenia, which have overlapping clinical features and are common initial misdiagnoses or prodromes of bvFTD, to extend the literature, and to potentially support clinical translation.

**<u>Aim 2</u>** was to use a comprehensive and sophisticated model of percentiles of NfL across the lifespan based on a large reference range cohort of healthy controls, to interpret and compare NfL levels in primary psychiatric disorders and bvFTD. This model, Model 1, will be used to develop an interactive web-based application to allow visualisation of an individual’s NfL level compared to this large reference cohort, outputting calculated individualised centiles and z-scores. This would facilitate future research relevant to diagnostic and clinical utility, and provide a potential clinical tool to interpret an individual patient’s NfL level at the bedside.

**<u>Aim 3</u>** was to investigate plasma NfL levels in a range of PPDs, compared to each other, and compared to controls. Most previous studies that focused on PPDs, did not include weight or BMI as covariate, which has been shown to be inversely associated with blood NfL levels (Benkert et al., 2022; Eratne et al., 2021; Manouchehrinia et al., 2020). Therefore, as secondary additional aims, the impact of weight as a covariate was explored with a model that did not include weight (Model 2), and a model that included weight (Model 3), in turn extending on Aim 2 and Model 1 to determine how sensitive comparisons/analyses to weight and when using different control groups, important once again for future research and clinical translation.

## METHODS

### Participant recruitment and data

Participant samples and data were included from four patient cohorts and two control groups, detailed below.

#### Cohort 1, bipolar affective disorder (BPAD)

Baseline (pre-intervention) samples and data were collected during a 16-week, three-arm, double-blind, randomised control trial (RCT) of adjunctive mitochondrial agents and *N*-acetylcysteine for bipolar depression [ACTRN12612000830897] (Berk et al., 2019). Participants were at least 18 years old, met DSM-IV-TR diagnostic criteria for bipolar disorder (assessed via structured clinical interview), and experiencing a bipolar depressive episode of at least moderate severity (n=121). Participants who were under any form of therapy needed to remain on stable treatment for at least 1 month prior to entering the study. Recruitment occurred between 2013 and 2015.

#### Cohort 2, major depressive disorder (MDD)

Baseline (pre-intervention) samples and data were collected during a 12-week, two-arm, double-blind RCT of adjunctive minocycline for unipolar depression [ACTRN12612000283875] (Dean et al., 2014, 2017). Participants were at least 18 years old, met DSM-IV diagnostic criteria for unipolar depression (assessed via structured clinical interview), and experiencing a current depressive episode of at least moderate severity (n=42). Participants currently under any therapy needed to remain on stable treatment for at least 2 weeks prior entering the study. Recruitment occurred between 2013 and 2015.

#### Cohort 3, treatment-resistant schizophrenia (TRS)

Participants were from the Cooperative Research Centre (CRC) Psychosis Study, a cross-sectional study that recruited people aged 18-65 years from inpatient and outpatient services in Melbourne, Australia, between 2012-2017 (n=82), who were on clozapine and had a diagnosis of treatment-resistant schizophrenia (TRS), defined as failure to respond to adequate trials of two or more antipsychotics, as previously described (Bousman et al., 2019; Eratne et al., 2021; Mostaid et al., 2017) The TRS biobank is detailed here: https://researchdata.ands.org.au/treatment-resistant-schizophrenia-biobank/1325206

#### Cohort 4, behavioural variant frontotemporal dementia (bvFTD)

Twenty-two patients were recruited from the Eastern Cognitive Disorders Clinic, Eastern Health, Melbourne, Australia, a specialist cognitive neurology service with expertise in diagnosis and management of bvFTD. Included in this study were patients who met diagnostic criteria for probable or definite bvFTD based on comprehensive gold-standard expert multidisciplinary and multimodal investigations including structural and functional imaging, as previously described (Ooi et al., 2022).

#### Control Group 1, local control group

Samples and data were pooled from healthy people (n=96) with no current or past psychiatric or neurological illness, from the CRC Psychosis Study (healthy controls age-matched to TRS, and healthy parents and siblings of participants with TRS), as described previously (Eratne et al., 2021).

#### Control Group 2, large reference normative control group

Data was available from 1,926 people aged 5-90 years, with no history or clinical symptoms or signs of neurological disorder, pooled from eight cohorts and as described previously in detail (Simrén et al., 2022). This data was used as a large reference population for modelling of NfL levels across most of the lifespan.

Common data from all studies included diagnosis, age, sex, and weight (where available). For Cohorts 3 and 4, and Control Groups 1 and 2, there was no: significant renal impairment, severe/uncontrolled diabetes or other general medical conditions, and no known stroke, head injury, within at least 12 months of recruitment. This data was not available for Cohorts 1 and 2. More detailed information on recruitment and eligibility criteria have been published previously (Berk et al., 2019; Bousman et al., 2019; Dean et al., 2014, 2017; Eratne et al., 2021; Mostaid et al., 2017; Ooi et al., 2022; Simrén et al., 2022).

All studies that contributed cohort data and samples to this study, had ethical approval at the relevant Human Research Ethics Committees, and all participants provided written informed consent prior to participation. This study, which is part of The Markers in Neuropsychiatric Disorders Study (The MiND Study, https://themindstudy.org), was approved by the Melbourne Health Human Research Ethics Committee (MH HREC 2020.142).

### Sample analysis

Plasma aliquots from all samples were stored at -80°C. For Cohorts 1-4 and Control Group 1 samples were randomised before analysis, and analyses were blinded to diagnosis. All plasma NfL levels were measured on the Quanterix SR-X and HD-X analysers using Simoa NF-Light kits, according to the manufacturer’s recommendations (Quanterix Corporation, Billerica, MA USA).

### Statistical analysis

All statistical analyses were performed using R version 4.2.2 (2022-10-31). For Aim 1 and Aim 3, general linear models (GLMs) were used to examine relationships between NfL levels, diagnostic group, and relevant clinicodemographic variables. For these models, log10-transformed NfL was entered as the dependent variable. Diagnostic group (using Control Group 1 as the reference class), age, sex, and weight (where available) were included as independent variables. For Aim 3, GLMs were performed with and without adjusting for weight to investigate the contribution of this covariate to overall results. 95% confidence intervals were computed for all GLMs via nonparametric bootstrapping (1000 replicates), with statistical significance defined as any confidence interval not including the null (at the 95% level). Receiver operator characteristic (ROC) curves were computed to estimate area under the curve (AUC), sensitivity, and specificity of NfL in distinguishing between groups. Optimal cut-off was determined using Youden’s method. For Aim 2, all patient cohorts and Control Group 1 were compared to the large reference cohort, Control Group 2. Z-scores were calculated from age-adjusted percentiles from Control Group 2, which were derived using generalised additive models for location, scale, and shape (GAMLSS). Single-sample t-tests were used to test the null hypothesis that the mean z-score was 0 (i.e., no difference / equal to the mean of Control Group 2). Welch Two Sample two-tests were used to compare z-scores between groups. The GAMLSS model was used to develop the web-based application. Model residuals were inspected for normality and homoscedasticity.

## RESULTS

The study included 245 participants with psychiatric disorders: 121 BPAD, 42 MDD, 82 TRS, ranging from 20 to 79 years of age (Table 1), 22 participants with bvFTD (mean age 66 years, range 43-80), 96 participants in Control Group 1 (mean age 45 years, range 18-77), and 1,926 participants in Control Group 2 (mean age 54 years, range 5-90). The bvFTD group was the oldest group (mean 66 years). MDD patients were older (mean 55 years) than BPAD and TRS (44 and 40 years, respectively). TRS and bvFTD had lower proportion of females (28% and 18%, respectively), compared to the other groups (all above 50%). Weight data was not available for two groups: bvFTD, and Control Group 2. Participants with TRS, all of whom were on clozapine, were heavier (mean weight 95.8kg) compared to the other psychiatric groups (BPAD 84.3kg, MDD 84.6kg) and Control Group 1 (77.3kg), with the most likely explanation being the common adverse effect of significant weight gain with clozapine.

**Table 1.** Study demographics and plasma neurofilament light levels. bvFTD: behavioural variant frontotemporal dementia. Data presented are mean [bootstrapped 95% confidence interval] or number (%).

|  | Bipolar<br>affective<br>disorder<br>(BPAD) | Major<br>depressive<br>disorder (MDD) | Treatment-resistant<br>schizophrenia<br>(TRS) | bvFTD | Control Group 1 | Control Group 2 |
| --- | --- | --- | --- | --- | --- | --- |
| N | 121 | 42 | 82 | 22 | 96 | 1926 |
| Age at sample, y | 44.1 [42.0,<br>46.3] | 55.0 [51.1,<br>58.9] | 40.3 [38.4,<br>42.4] | 65.7 [61.6, 69.7] | 44.7 [41.8, 47.4] | 54.4 [53.8, 55.0] |
| Age range, y | 20-72 | 26-79 | 22-61 | 43-80 | 20-77 | 5-90 |
| Sex, n female (%) | 75 (62%) | 24 (57%) | 23 (28%) | 4 (18%) | 50 (52%) | 1218 (63%) |
| Weight, kg | 84.3 [81.0,<br>87.8] | 84.6 [79.3,<br>89.8] | 95.8 [90.2,<br>102.3] (n=71) | - | 77.3 [74.1, 80.7]<br>(n=87) | - |
| Plasma NfL, pg/mL | 8.4 [7.6, 9.4] | 10.9 [9.1, 13.9] | 6.6 [5.8, 7.9] | 44.5 [33.0, 64.5] | 9.4 [7.3, 18.2] | 9.9 [9.6, 10.2] |
| Log10NfL | 0.87 [0.83,<br>0.91] | 0.97 [0.90,<br>1.00] | 0.73 [0.67,<br>0.79] | 1.54 [1.38, 1.68] | 0.85 [0.80, 0.91] | 0.93 [0.91, 0.94] |
| z-scores (Model 3) | 0.44 [0.23, 0<br>64] | 0.26 [-0.04,<br>0.57] | -0.10 [-0.39,<br>0.19] | 2.02 [1.32, 2.58] | 0.18 [-0.06, 0.43] | 0 (reference group) |

### Aim 1: Plasma NfL in behavioural variant frontotemporal dementia compared to primary psychiatric disorders

As demonstrated in Table 1 and Figure 1, the highest plasma NfL levels were seen in bvFTD (mean NfL, M=44.5pg/mL, 95% confidence interval CI [33.0, 64.5]) and were approximately four times higher than all other groups (all mean levels<11pg/mL).

**Figure 1.**
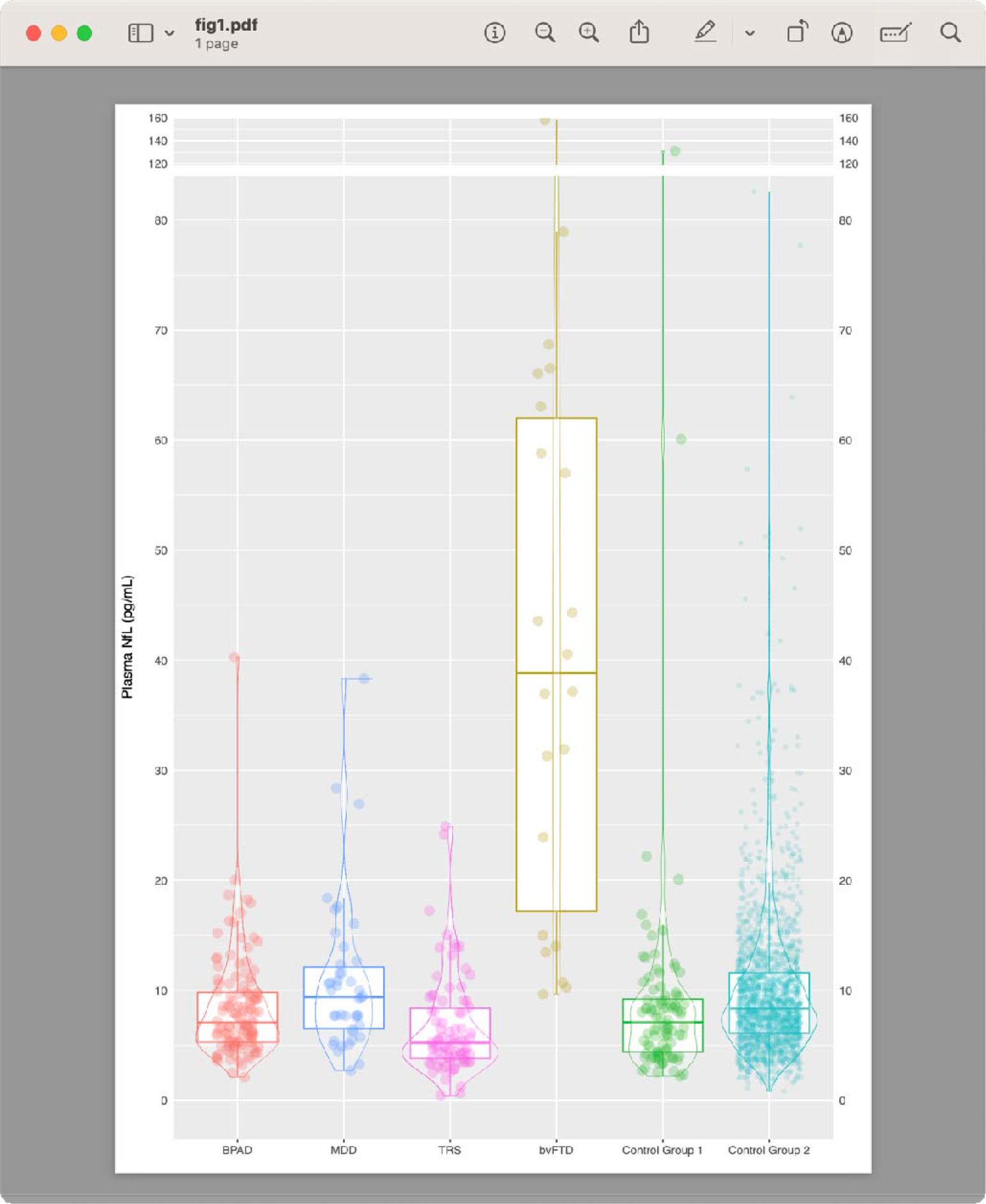
Plasma NfL levels in primary psychiatric disorders, behavioural variant frontotemporal dementia, and controls BPAD: bipolar disorder; bvFTD: behavioural variant frontotemporal dementia; MDD: major depressive disorder; TRS: treatment-resistant schizophrenia

A GLM to compare log NfL levels in bvFTD to all other groups, adjusting for age and sex, demonstrated statistically significant and large differences: BPAD (β MDD (β=1.80 [1.09, 2.39], p<0.001), TRS (β=1.98 [1.36, 2.62], p<0.001), and Control Group 1 (β =1.94 [1.35, 2.51], p<0.001).

ROC curve analyses were performed to assess whether plasma NfL could distinguish bvFTD from other groups. Plasma NfL distinguished between bvFTD and all psychiatric disorders, with high accuracy (AUC=0.95 [0.91, 0.99]). An optimal cut-off of 13.3pg/mL resulted in 86% specificity, 88% sensitivity). Diagnostic performance remained high even when restricting psychiatric disorders to the same age range as the bvFTD group (43-80 years): AUC 0.91 [0.85, 0.98], 13.3pg/mL cut-off, 86% specificity, 78% sensitivity (and a cut-off of 22 resulted in 73% specificity, but higher sensitivity 95%) Additional details are available in the Supplementary Material Table 1.

### Aim 2: Plasma NfL in all groups compared to large reference normative control cohort (GAMLSS model, Model 1)

To explore the utility of a large normative control dataset, age-adjusted percentiles for plasma NfL were derived from the reference cohort Control Group 2, using GAMLSS. The centile plot of this model (Model 1) across most of the lifespan, is demonstrated in Figure 2.

**Figure 2.**
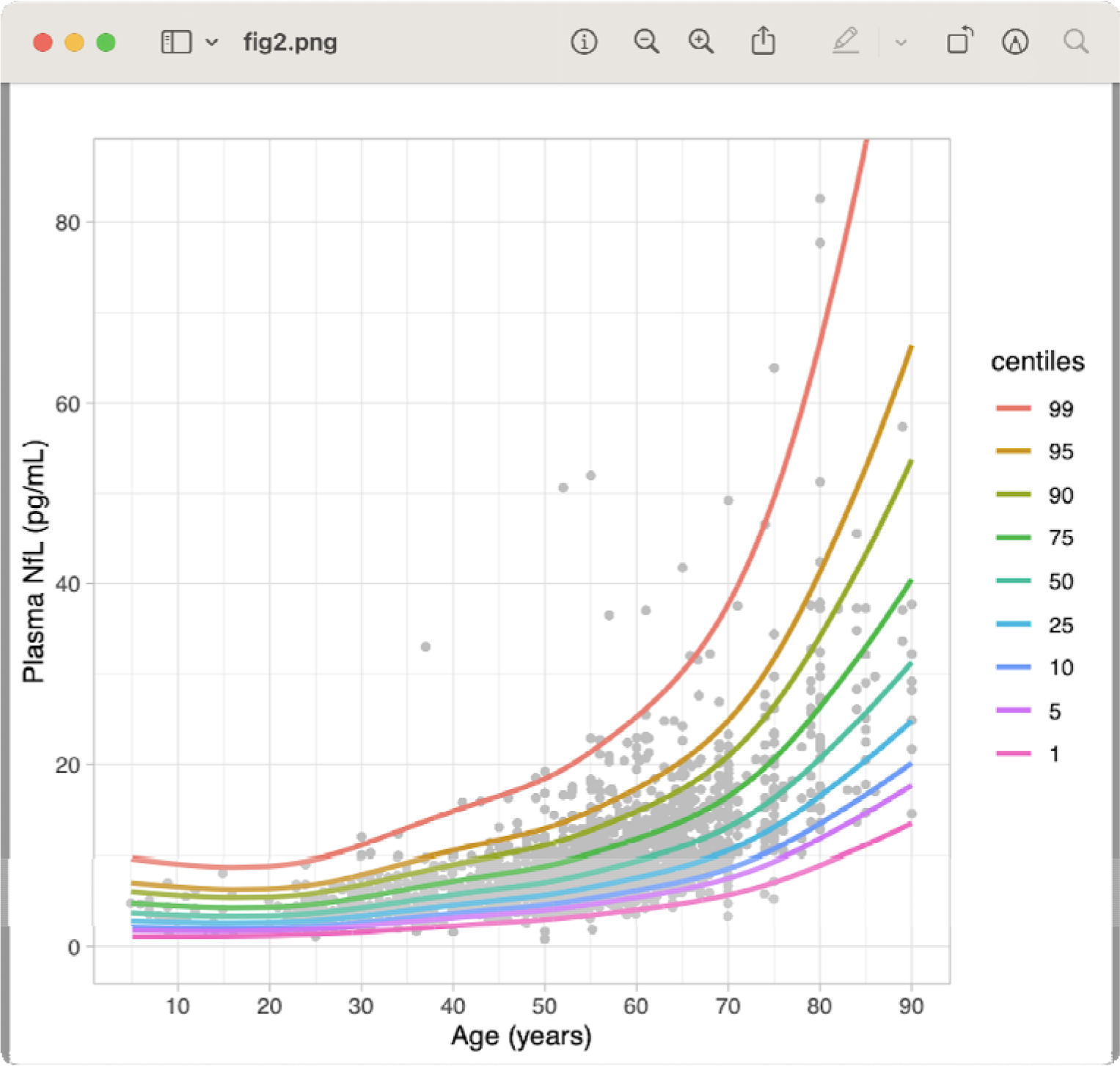
Percentiles derived from generalised additive models for location, scale, and shape, from 1926 healthy controls, Control Group 2 (Simrén et al., 2022)

To investigate any differences, z-scores for individuals in every cohort were computed from this reference cohort model. As demonstrated in Figure 3, highest z-scores were in bvFTD (M=2.02 [1.32, 2.58]), compared to all other groups. bvFTD was significantly greater than Control Group 2, and the effect was large (difference=2.02 [1.41, 2.62], p < .001; Cohen’s d=1.47 [0.85, 2.07]). bvFTD was greater than BPAD, MDD, TRS, Control Group 1 (differences=1.58, 1.75, 2.02, 1.84, respectively), and all with large effect sizes (Cohen’s d=1.25 [1.48, 1.58, 1.43], respectively).

**Figure 3.**
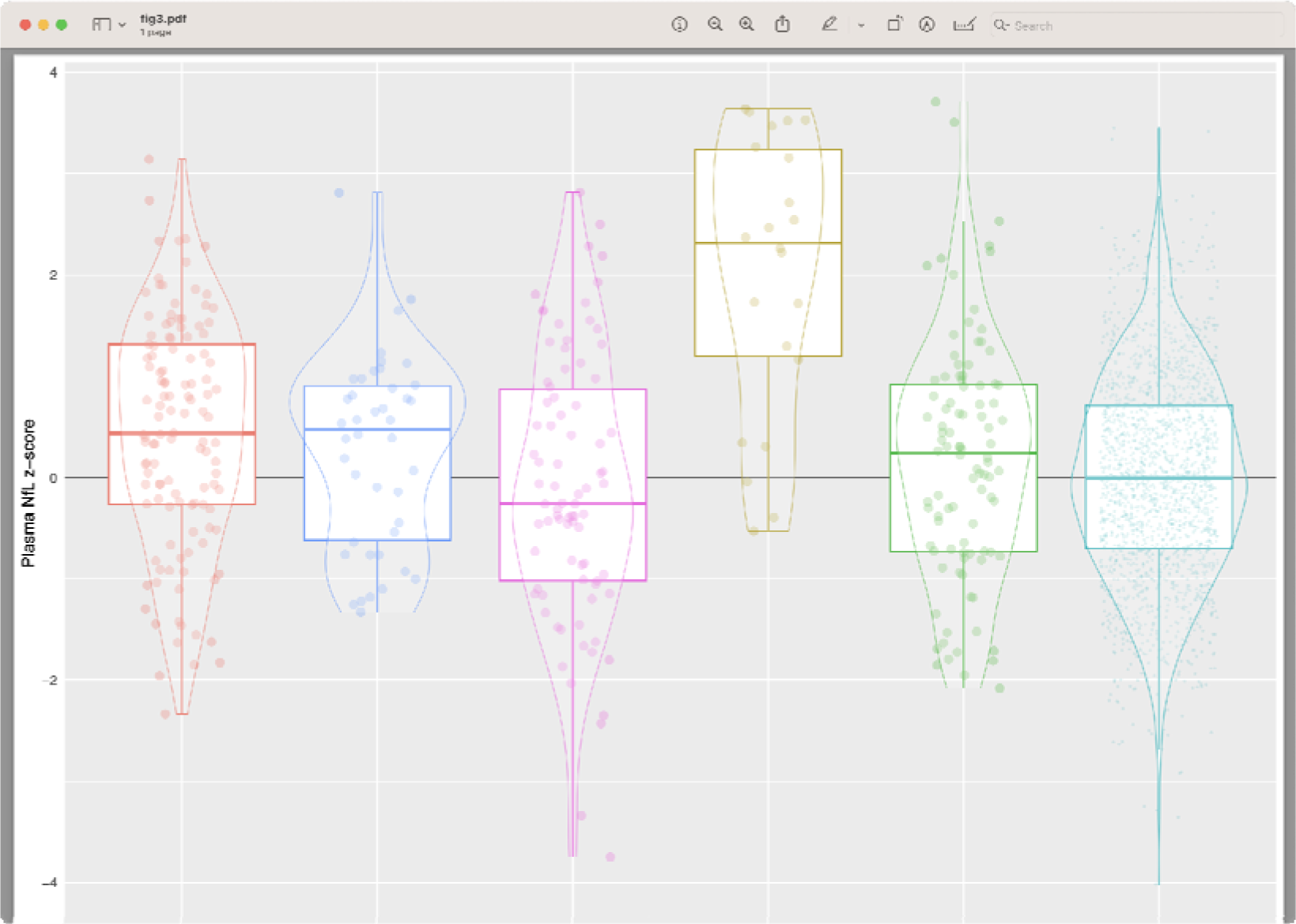
Z-scores (compared to Control Group 2) of plasma NfL levels in primary psychiatric disorders, behavioural variant frontotemporal dementia, and controls BPAD: bipolar disorder; bvFTD: behavioural variant frontotemporal dementia; MDD: major depressive disorder; TRS: treatment-resistant schizophrenia

Focusing on PPDs, the mean z-score for BPAD was greater than Control Group 2, suggesting a small effect (mean z-score 0.44 vs 0, difference=0.44 [0.23, 0.64], p<0.001; Cohen’s d=0.39 [0.20, 0.57]).

BPAD was also greater than TRS, again suggesting a small effect (mean z-score 0.44 vs -0.10, difference=0.54 [0.19, 0.90], p=0.003; small effect Cohen’s d=0.44 [0.15, 0.73]). Z-scores in the other groups (MDD, TRS, and Control Group 1), were not different to Control Group 2.

### Aim 3: Plasma NfL in primary psychiatric disorders

Before adjusting for age, raw NfL in psychiatric disorders were highest in MDD (mean M=10.9pg/mL 95%CI [9.1, 13.2], mean log10nfl=0.97 [0.90, 1.00]), with lowest levels in TRS (M=6.6pg/mL [5.8, 7.6]), log10nfl=0.73 [0.67, 0.79]). However, these were also the oldest and youngest groups, respectively (mean age 55 years vs 40). Therefore GLMs were performed to control for these age differences and derive adjusted/estimated marginal means, to compare differences between psychiatric disorders and controls, as detailed below (full details are also available in Supplementary Material). For exploratory analyses on the impact of weight as a covariate, two GLMs were performed: one without weight (Model 2), and one including weight (Model 3).

#### Model 2 (GLM without weight)

A GLM adjusting for age and sex (without weight) was used to compare mean log NfL differences between psychiatric disorders and Control Group 1. No differences were seen between BPAD and Control Group 1 (estimated marginal mean log10NfL, EMM, 0.98 95%CI:[0.93, 1.00] vs 0.95 [0.91, 0.98], p=0.24), and MDD and Control Group 1 (EMM 0.94 [0.89, 1.00] vs 0.95 [0.91, 0.98]), p=0.83). In this model, TRS had lower levels than Control Group 1 (EMM 0.89 [0.85, 0.93] vs 0.95 [0.91, 0.98], p=0.030). Comparing psychiatric groups to each other, levels were not different between MDD and BPAD (EMM 0.94 [0.89, 1.00] vs 0.98 [0.93, 1.00], p=0.41). Levels were also lower in TRS compared to BPAD (EMM 0.89 [0.85, 0.93] vs 0.98 [0.93, 1.00], p=0.004) and TRS compared to MDD (EMM 0.89 [0.85, 0.93] vs 0.94 [0.89, 1.00], p=0.042).

#### Model 3 (GLM including weight)

Adding weight to the GLM for Model 3, resulted in no significant differences between TRS and MDD (EMM 0.80 [0.75, 0.84] vs 0.85 [0.78, 0.86], p=0.20), and between TRS and Control Group 1 (EMM 0.80 [0.75, 0.84] vs 0.82 [0.78, 0.86], p=0.45), different to results from Model 2. Including weight showed statistically higher NfL levels in BPAD compared to Control Group 1 (EMM 0.88 [0.84, 0.91] vs 0.82 [0.78, 0.86], p=0.028). Levels remained statistically significantly lower in TRS compared to BPAD (EMM 0.80 [0.75, 0.84] vs 0.88 [0.84, 0.91], p=0.020).

Weight had a small effect (β =0.04 [-0.14, 0.22], p=0.640), but was in Model 3 (β=0.19 [0.02, 0.36], p=0.028). Sensitivity analyses were performed, excluding sex as a covariate, finding no impact on the overall findings. As expected, age had the highest coefficients in both Models 2 and 3 (β =0.61 [0.54, 0.68], p<0.001, respectively).

ROC curve analyses demonstrated that although these statistically significant differences were found in Models 2 and 3, plasma NfL did not demonstrate high diagnostic utility to distinguish between TRS and BPAD (AUC 0.66 [0.58, 0.74], 5.6pg/mL cut-off, 72% sensitivity, 59% specificity), TRS and MDD (AUC 0.74 [0.65, 0.83], 6.2pg/mL cut-off, 79% sensitivity, 63% specificity). Plasma NfL did not accurately distinguish BPAD from Control Group 1 (AUC 0.46 [0.38, 0.53]).

Of note, Model 1 (z-scores from a large reference control group, without weight), resulted in similar findings to the model comparing to a smaller local control group, and including weight (Model 3) – i.e. Model 1, like Model 3, found differences between BPAD and Controls, and between BPAD and TRS.

## DISCUSSION

There were three main findings from this study. First, significantly higher plasma NfL levels were seen in bvFTD compared to primary psychiatric disorders and controls, and high plasma NfL demonstrated high diagnostic accuracy in distinguishing bvFTD from PPD. Second, GAMLSS modelling and z-scores using a large reference control group, performed just as well as models using local controls that included weight, and outperformed models that did not include weight, while also allowing for more precise interpretation and visualisation of individual levels by facilitating z-score and percentiles across the lifespan. Finally, slightly higher group NfL levels were found in patients with BPAD compared to controls, and in BPAD compared to patients with treatment resistant schizophrenia.

Finding elevated plasma NfL levels in bvFTD compared to a large group of well-characterised PPD (including to our knowledge, the largest blood NfL BPAD cohort), extends the previous literature. The strong diagnostic performance of plasma NfL to distinguish bvFTD from PPD, supports the clinical use of a relatively easily accessible blood biomarker, to assist in this challenging, clinical distinction (Al Shweiki et al., 2019; Ducharme et al., 2020; Eratne, Keem, et al., 2022; Katisko et al., 2020; Ooi et al., 2022), of high relevance to a diverse range of clinicians and clinical settings.

The robust performance of Model 1, using a large reference cohort for comparisons (Simrén et al., 2022), equalling or outperforming other models using locally recruited controls and including weight as a covariate, has important potential implications. Not only could there be significant improvements in accuracy of comparisons and interpretations for future research and clinical purposes, but significant efficiencies and cost reductions may be possible for future studies by potentially not requiring local control group recruitment, and instead focusing on facilitating data sharing and pooling. Z-scores provide useful information on the degree of deviation and therefore strength/confidence in interpreting an individual level, allowing for a more precision interpretation, and mitigating some of the limitations of age-binned cut-offs and coarse interpretation binary of ‘normal’ or ‘abnormal’ based on being above or below the cut-off, especially given the non-linear relationship and increasingly sharp change in slopes of NfL with age (as demonstrated in Figures 2). Based on Model 1, we developed an interactive web-based application, available via https://themindstudy.org/apps. The application allows input of an individual’s age and plasma NfL level (generated using technology/assays similar to those used for the reference cohort), providing estimated centiles and z-scores and allowing visualisation on the centiles reference chart, compared to the large reference cohort, Control Group 2. This builds on other similar applications that have been developed recently (Benkert et al., 2022; Vermunt et al., 2022), but to our knowledge this is the first application to use GAMLSS modelling and providing both individualised percentiles and z-scores for plasma NfL. This application could be used for academic and research interests and will be used in studies underway to investigate the clinical and diagnostic utility and validity of such tools, as well as feasibility and utility for clinicians. One potential implementation in routine clinical care in the future, could be where clinician uses such an application, akin to using growth charts, to help quickly facilitate a precision interpretation of an individual’s NfL level (as demonstrated with examples in Figure 4).

**Figure 4.**
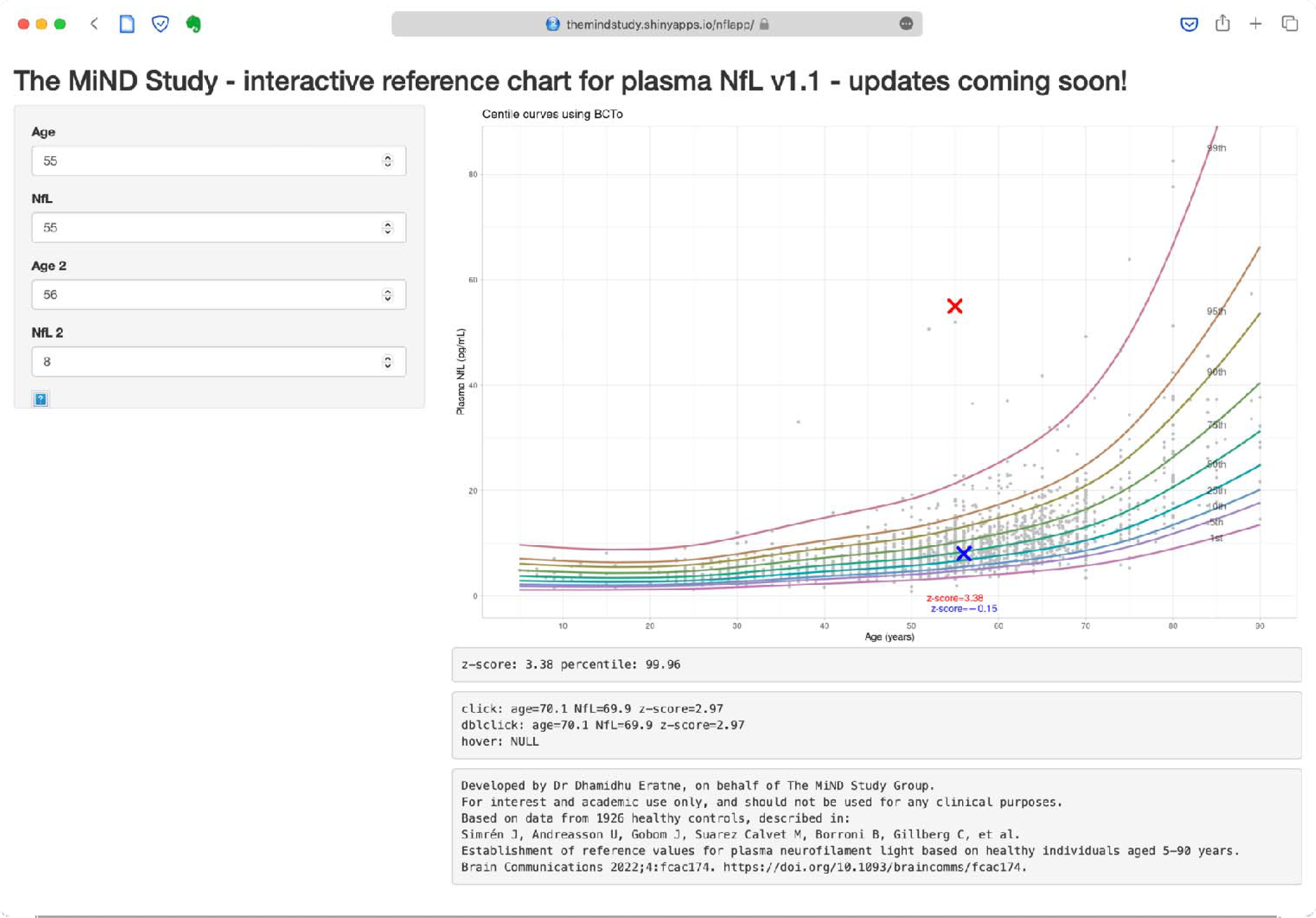
The interactive web-based application The user can input individual ages and plasma NfL levels, providing estimated centiles and z-scores and allowing visualisation on the centiles reference chart, compared to the large reference cohort, Control Group 2. The application is available via https://themindstudy.org/apps. Two examples have been entered (real patients, not part of the study cohort). The red cross refers to patient 1, a 55-year-old man who was initially diagnosed with late onset psychosis and delirium, but on a third subsequent reassessment he was eventually re-diagnosed with probable behavioural variant frontotemporal dementia (bvFTD), and ultimately definite bvFTD (genetic testing confirmed the *C9orf72* repeat expansion mutation). In hindsight the psychosis diagnosis was very likely a prodrome/misdiagnosis. His plasma NfL level was 55pg/mL, which as the figure conveys visually was significantly elevated (99.96^th^ percentile and z-score of 3.38 for a 55-year-old), was consistent with a neurodegenerative disorder and could have quickly dismissed primary psychiatci and non-neurodegenerative differential diagnoses. Conversely, patient 2’s level of 8pg/mL (blue cross, z-score of 0.15) could have potentially dismissed or at the very least added caution to an initial diagnosis of probable bvFTD, which in hindsight was a misdiagnosis and this 56-year-old woman was eventually re-diagnosed to bipolar disorder.

We explored the sensitivity of using different control groups and the influence of weight as a covariate. We found that Model 1, using the large control reference data set without weight, had the same overall results as analyses from specifically recruited local controls and when weight was included as a covariate (Model 3). Of note, Model 1 avoided some of the spurious findings of a model using local controls that did not include weight (i.e., Model 2, which suggested lower levels in TRS compared to controls and MDD). The finding that weight may not be required, when using a large control data set and using GAMLSS modelling such as in Model 1, requires further investigation, but has potential important implications. While there is growing evidence of weight and other factors that influence plasma NfL levels, the overall impact of these is relatively small (Akamine et al., 2020; Fitzgerald et al., 2022). Considering clinical translation, a clinician only having to consider and input only three simple and easily/immediately obtainable variables for many patients – age, sex, and plasma NfL level – would be more feasible for busy clinicians in primary care and via telehealth/remote assessments, and even has implications for laboratories reporting on plasma NfL levels, while reducing the potential for variability introduced by weight measurements in different clinical and research settings. An important limitation, however, was the lack of weight data in all groups, and therefore to further investigate and extend our findings, future studies should incorporate as many of these variables into analyses and modelling, and to specifically investigate the clinical utility of including these variables, compared to just a minimum set (e.g., age, sex, plasma NfL).

We identified statistically higher NfL levels in BPAD, compared to controls, and compared to TRS. This is in the largest group of BPAD in whom to date, to our knowledge, blood NfL levels have been described. A previous study found elevated serum NfL levels in 45 people with bipolar depression (Aggio et al., 2022), although the control group was not age-matched and the difference between raw/unadjusted mean levels between BPAD and controls was much larger in that study (9.13pg/mL vs 4.28pg/mL), compared to our study (8.4pg/mL in BPAD, vs 9.4 and 9.9pg/mL in Control Groups 1 and 2, respectively). Elevated CSF NfL levels in BPAD have previously been described (Jakobsson et al., 2014; Rolstad et al., 2015). Our finding of elevated levels in BPAD compared to controls may suggest some degree of mild and/or slow rate of neuronal injury in BPAD, greater than in controls/healthy ageing, but less than what is seen in clearly neurodegenerative disorders such as bvFTD and Alzheimer’s disease (Ashton et al., 2021). To further extend these findings in BPAD, future studies with serial NfL levels, associations with additional biomarkers (e.g., of inflammation/neuroinflammation) and neuroimaging, medications/treatments, different stages/phases of illness (e.g., bipolar mania), and longitudinal comprehensive follow up, will be valuable. It is important to note that while there were statistically significant differences between BPAD and control and TRS groups, the differences and effects were small, there was significant overlap between groups, and correspondingly poor performance on ROC curve analyses. Therefore, plasma NfL to distinguish BPAD from other primary psychiatric conditions has no clinical utility, while the diagnostic utility to distinguish BPAD from bvFTD, remains high.

This study demonstrated the strong diagnostic utility of plasma NfL in distinguishing bvFTD from clinically relevant PPDs, building the accumulating evidence base for a simple blood test to assist with this common yet challenging diagnostic dilemma, of high relevance to a broad range of clinicians from primary care to neurology, psychiatric, geriatric and memory clinic settings. In addition, this study found no significant differences between MDD, TRS, and controls, and small elevations in BPAD compared to controls, adding to our understanding of these disorders and evidence suggesting a lack of significant neuronal injury and degeneration (axonal in particular) in primary PPDs. The application developed demonstrates how visualisation of an individual’s NfL level and minimal additional data (age), using a large reference cohort and sophisticated modelling, moves beyond coarser age-based binary cut-offs and starts to facilitate the individualised and precise medicine interpretation required for best translation into real-world clinical care. Studies are underway to investigate the clinical and diagnostic utility of plasma NfL and the application, in diverse neurodegenerative and primary psychiatric conditions in real-world primary care, and specialist clinical settings.

## Data Availability

All data produced in the present work are contained in the manuscript

## ACKNOWLEDGEMENTS AND FUNDING SOURCES

The authors acknowledge the financial support of the CRC for Mental Health. The Cooperative Research Centre (CRC) programme is an Australian Government Initiative. The authors wish to acknowledge the CRC Scientific Advisory Committee, in addition to the contributions of study participants, clinicians at recruitment services, staff at the Murdoch Children’s Research Institute, staff at the Australian Imaging, Biomarkers and Lifestyle Flagship Study ofAging, and research staff at the Melbourne Neuropsychiatry Centre, including coordinators Merritt, A., Phassouliotis, C., and research assistants, Burnside, A., Cross, H., Gale, S., and Tahtalian, S. Participants for this study were sourced, in part, through the Australian Schizophrenia Research Bank (ASRB), which is supported by the National Health and Medical Research Council of Australia (Enabling Grant N. 386500), the Pratt Foundation, Ramsay Health Care, the Viertel Charitable Foundation and the Schizophrenia Research Institute. We thank the Chief Investigators and ASRB Manager: Carr, V., Schall, U., Scott, R., Jablensky, A., Mowry, B., Michie, P., Catts, S., Henskens, F., Pantelis, C., Loughland, C. We acknowledge the help of Jason Bridge for ASRB database queries. The authors are grateful for assistance from Brett Trounson and Dr Christopher Fowler and the team at The Florey Oak St Biobank.

AJW is supported by a Trisno Family Fellowship, funded in part by an NHMRC CRE (1153607). C Pantelis was supported by a National Health and Medical Research Council (NHMRC) Senior Principal Research Fellowship (1105825), an NHMRC L3 Investigator Grant (1196508).

HZ is a Wallenberg Scholar supported by grants from the Swedish Research Council (#2022-01018), the European Union’s Horizon Europe research and innovation programme under grant agreement No 101053962, Swedish State Support for Clinical Research (#ALFGBG-71320), the Alzheimer Drug Discovery Foundation (ADDF), USA (#201809-2016862), the AD Strategic Fund and the Alzheimer’s Association (#ADSF-21-831376-C, #ADSF-21-831381-C, and #ADSF-21-831377-C), the Bluefield

Project, the Olav Thon Foundation, the Erling-Persson Family Foundation, Stiftelsen för Gamla Tjänarinnor, Hjärnfonden, Sweden (#FO2022-0270), the European Union’s Horizon 2020 research and innovation programme under the Marie Skłodowska-Curie grant agreement No 860197 (MIRIADE), the European Union Joint Programme – Neurodegenerative Disease Research (JPND2021-00694), and the UK Dementia Research Institute at UCL (UKDRI-1003).

KB is supported by the Swedish Research Council (#2017-00915 and #2022-00732), the Swedish Alzheimer Foundation (#AF-930351, #AF-939721 and #AF-968270), Hjärnfonden, Sweden (#FO2017-0243 and #ALZ2022-0006), the Swedish state under the agreement between the Swedish government and the County Councils, the ALF-agreement (#ALFGBG-715986 and #ALFGBG- 965240), the European Union Joint Program for Neurodegenerative Disorders (JPND2019-466-236), the Alzheimer’s Association 2021 Zenith Award (ZEN-21-848495), and the Alzheimer’s Association 2022-2025 Grant (SG-23-1038904 QC).

This study was supported by was also supported by: MACH MRFF RART 2.2, NHMRC (1185180), and Psychiatry and Rehabilitation Division, Region Skåne, Sweden. The role of these funding sources was to support research study staff and biosample analyses.

Finally, the authors would like to thank all the participants and their families for their participation.

The corresponding author had full access to all the data in the study and had final responsibility for the decision to submit for publication.

## Statistical analysis conducted by

Dr Dhamidhu Eratne A/Prof Charles Malpas Dr Steve Simpson-Yap

## SUPPLEMENTARY MATERIAL

Additional information on bvFTD vs all other group comparisons

**Supplementary Table 1.**
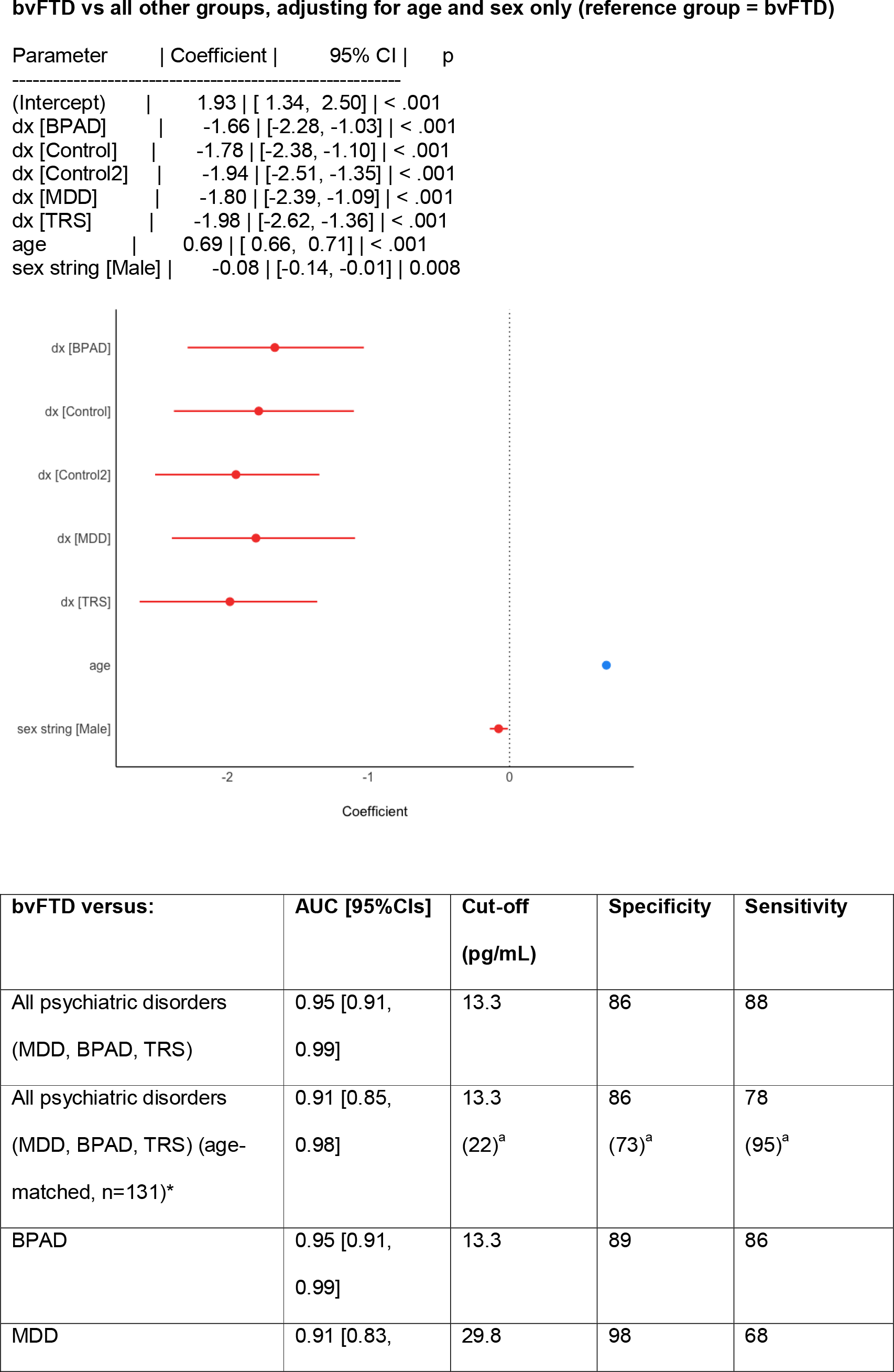
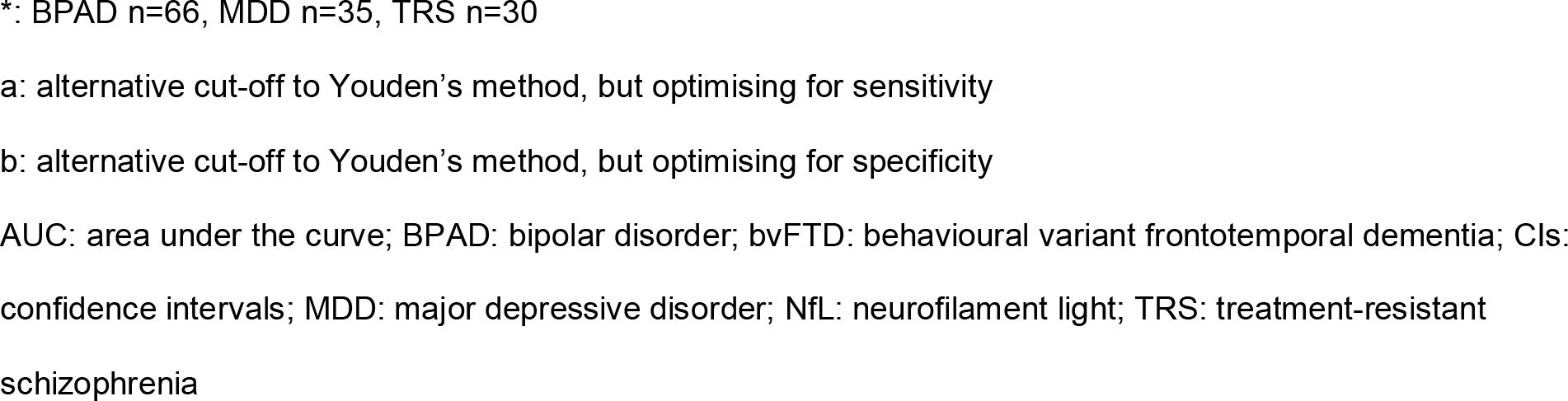
Receiver operating characteristic (ROC) curve analyses for bvFTD versus other clinically relevant groups. Levels above the cut-off indicate bvFTD. Results were similar when restricted to age range of bvFTD (43-80 years of age).

**Figure 1.**
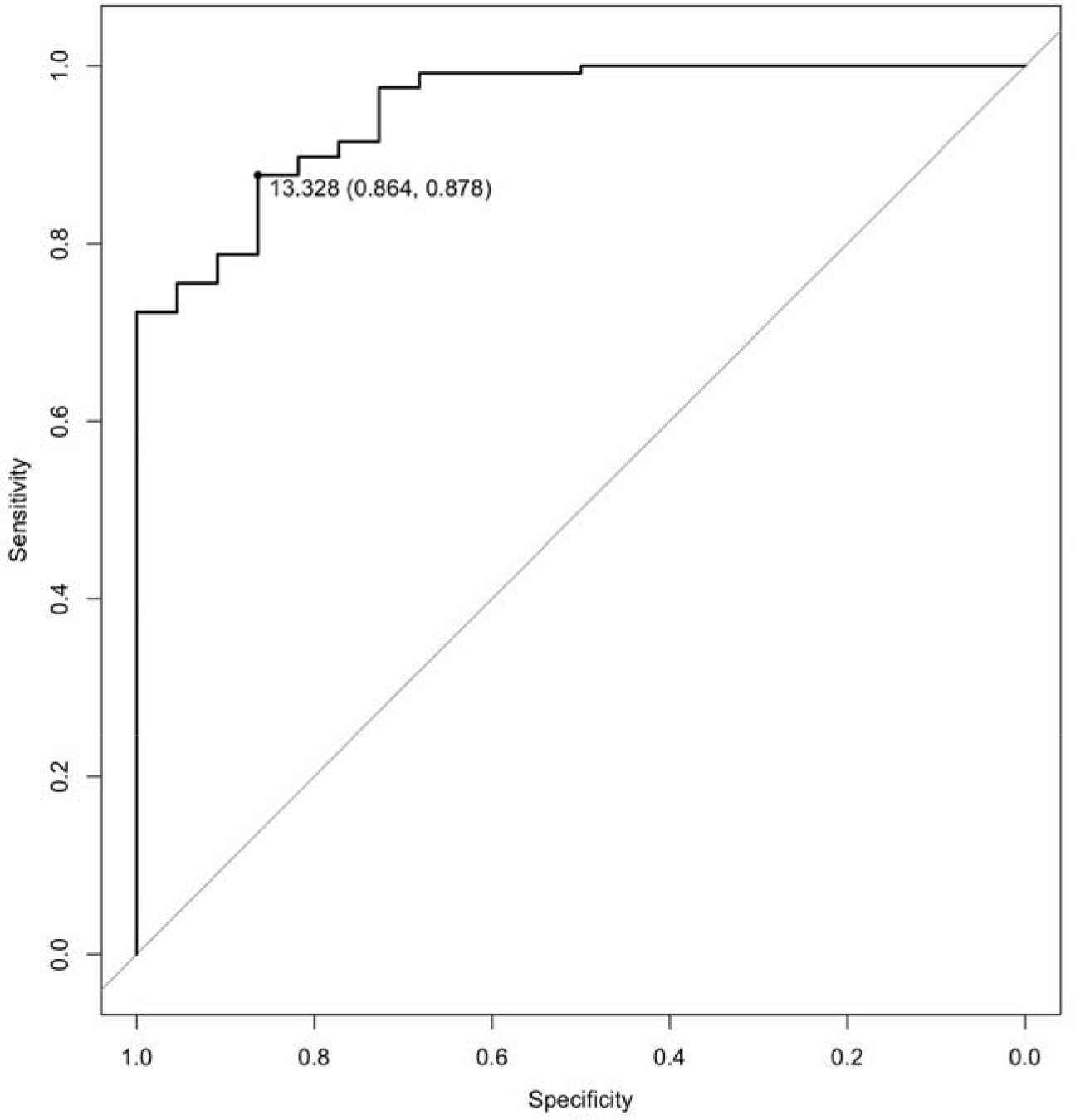
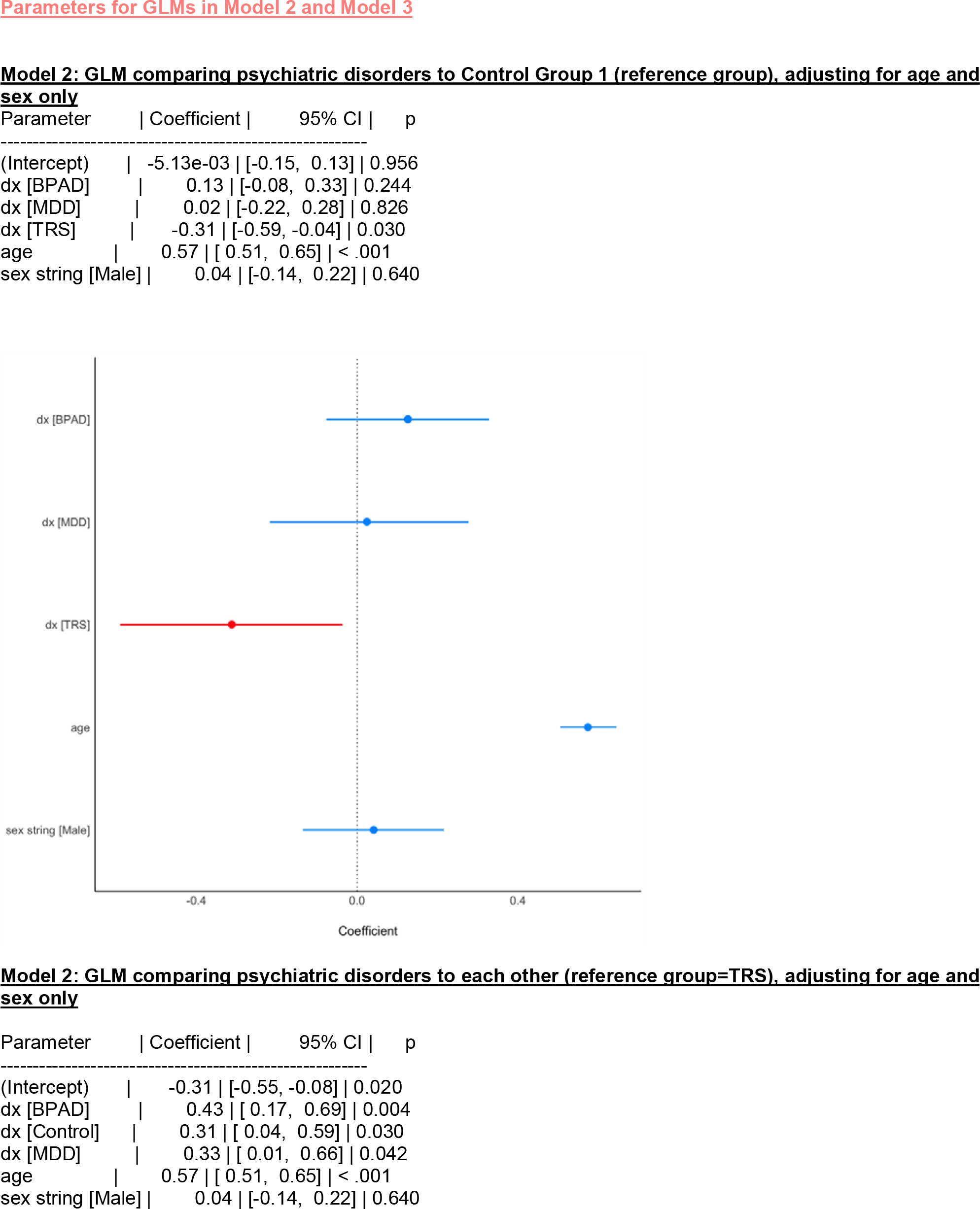
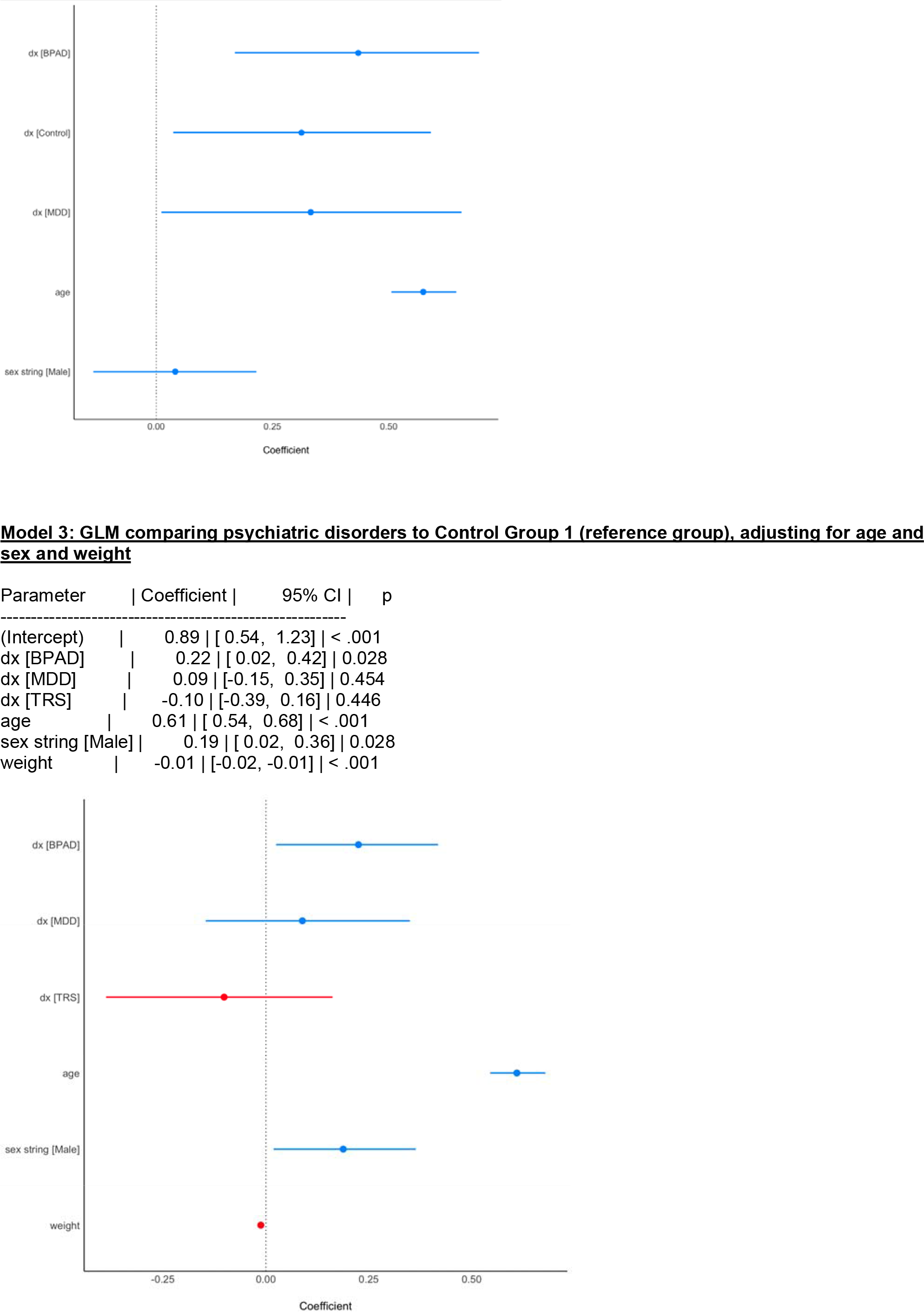
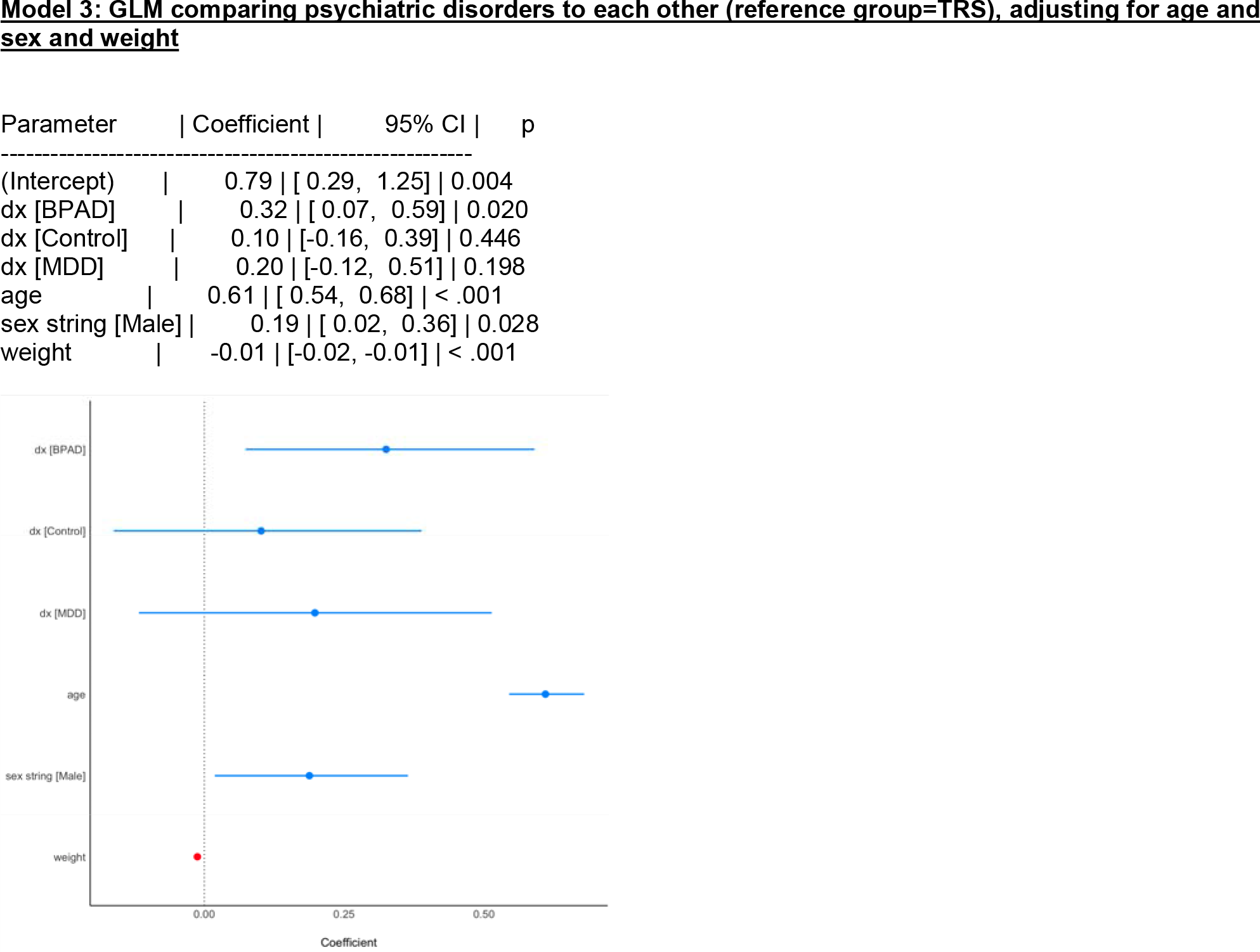

